# Evaluating Vision and Pathology Foundation Models for Computational Pathology: A Comprehensive Benchmark Study

**DOI:** 10.1101/2025.05.08.25327250

**Authors:** Rohan Bareja, Francisco Carrillo-Perez, Yuanning Zheng, Marija Pizurica, Tarak Nath Nandi, Jeanne Shen, Ravi Madduri, Olivier Gevaert

## Abstract

To advance precision medicine in pathology, robust AI-driven foundation models are increasingly needed to uncover complex patterns in large-scale pathology datasets, enabling more accurate disease detection, classification, and prognostic insights. However, despite substantial progress in deep learning and computer vision, the comparative performance and generalizability of these pathology foundation models across diverse histopathological datasets and tasks remain largely unexamined. In this study, we conduct a comprehensive benchmarking of 31 AI foundation models for computational pathology, including general vision models (VM), general vision-language models (VLM), pathology-specific vision models (Path-VM), and pathology-specific vision-language models (Path-VLM), evaluated over 41 tasks sourced from TCGA, CPTAC, external benchmarking datasets, and out-of-domain datasets. Our study demonstrates that Virchow2, a pathology foundation model, delivered the highest performance across TCGA, CPTAC, and external tasks, highlighting its effectiveness in diverse histopathological evaluations. We also show that Path-VM outperformed both Path-VLM and VM, securing top rankings across tasks despite lacking a statistically significant edge over vision models. Our findings reveal that model size and data size did not consistently correlate with improved performance in pathology foundation models, challenging assumptions about scaling in histopathological applications. Lastly, our study demonstrates that a fusion model, integrating top-performing foundation models, achieved superior generalization across external tasks and diverse tissues in histopathological analysis. These findings emphasize the need for further research to understand the underlying factors influencing model performance and to develop strategies that enhance the generalizability and robustness of pathology-specific vision foundation models across different tissue types and datasets.

## Introduction

Pathology plays a crucial role in cancer research, providing insights that drive the understanding, diagnosis, and treatment of cancer^1^. It involves thorough examination of tissue specimens for tumor diagnosis and identification of important prognostic features such as tumor stage and grade, as well as biomarkers suggestive of treatment response^2^. Pathologists study digital pathology slides to analyze tumor histologic features, including architectural patterns and cellular morphology, in order to detect and classify tumors^3^. Consequently, oncologic pathology plays an integral role in personalized medicine and treatment selection. Computational pathology (CPath) has enhanced the field of pathology by integrating automated workflows and processes and advanced computational techniques^1^. It has the potential to reduce the workload of pathologists considerably and can aid in speeding up the overall diagnostic process. With the advent of machine learning and artificial intelligence, CPath has proven potential to facilitate detection, classification and predictive modeling of cancer^4^. Further, CPath proved effective in assisting pathologists in tumor grading, staging, and subtyping^5^, while supporting diagnostic decision-making. Also, it has been employed as a computational technique to study patterns, segment tumors^6^, and extract intricate features, which enhances cancer detection^7^.

The use of Artificial Intelligence (AI) has been at the forefront of medical research for the past decade^8^. AI algorithms based on convolutional neural networks and vision transformers have significantly advanced computational pathology image analysis, especially in automated tumor detection and classification ^9^. AI-driven predictive models can analyze pathology images along with clinical and genomic data to predict patient outcomes, including disease progression, recurrence risk, and treatment response^10–12^. Integrating gene expression, spatial omics, single-cell omics, and digital pathology data with AI enables a comprehensive multi-modal approach to cancer diagnosis and biomarker discovery, leveraging deep learning to uncover intricate spatial and molecular relationships within the tumor microenvironment^13,14^. AI models can automatically segment and delineate tumor regions, extract complex features and perform risk based stratification^15^. Additionally, AI has accelerated Computational Pathology (CPath) by automating image analysis, streamlining workflows, integrating multimodal data^16,17^, and enhancing diagnostic accuracy^18^.

Despite these advancements, several challenges persist, particularly regarding data limitations and model generalizability. Pathology datasets suffer from shortage of labeled data as manual pathologist annotations are time-consuming and also cost ineffective^19^. In contrast, large-scale datasets like ImageNet^20^ benefit from extensive labeled data, enabling supervised learning approaches to achieve high accuracy across diverse natural image tasks. To address this, Self-Supervised Learning (SSL) has emerged as a machine learning paradigm where models are trained on large-scale datasets to learn meaningful representations without the need for labeled examples ^21^. This approach reduces the burden associated with data labeling and speeds up development of AI models. SSL based foundation models can be pre-trained on diverse and large unlabeled datasets,^22,23^ to learn generalized patterns, and then fine-tuned on specific pathology datasets of interest ^24^. SSL enables these foundation models to adapt to different types of pathologies, domains, and tasks, and improve their generalizability.^25,26^ Therefore there has been a large effort in developing general purpose pathology foundation models that are trained on millions of whole slide images^26–28^, and can be adapted to tumor classification, segmentation, grading tasks with smaller amounts of labeled data.

One of the first prominent models to be released was CTransPath^29^, which was pretrained on the TCGA^30^ and PAIP datasets^31,32^ and trained on approximately 15 million images. Lunit’s DINO-based ViT-Small model ^24^ also utilized TCGA, along with internal datasets comprising over 32.6 million patches and 36,000 slides. Phikon^33^, another SSL model based on the iBOT framework, was pretrained on 40 million images from TCGA, covering 16 cancer types. More recently, several models such as UNI^25^, Virchow^26^, and Hibou^27^ have adopted the DINOv2 SSL method with ViT-Large architectures for their pretraining, using proprietary datasets. Next, pathology language models such as PLIP ^34^, Quilt-Net ^35^, and CONCH ^36^, which are trained on both pathology slides and paired text, have demonstrated significant performance in vision-based tasks as well. Vision encoders like CLIP^37^, iBOT^23^ that are built into these models have shown competitive results in natural image classification and image retrieval tasks. Additionally, imagenet pre-trained models have been widely used as feature extractors for digital pathology.^38–40^

Despite these advancements in SSL, current foundation models may still be biased toward datasets they are trained on, limiting their transferability across diverse institutions and staining protocols^41^. Additionally, biases can arise from imbalanced representation of tissue types, and the predominance of certain cancer subtypes in training data, all of which can impact the model’s generalization and clinical applicability^42,43^. Many pathology foundation models rely on self-supervised pre-training using datasets like TCGA^24,33,44^ or large proprietary datasets from a single institution^25,26^, which can inadvertently result in domain-specific overfitting rather than true generalization across diverse clinical settings.^43^ This is further compounded by the limited availability of large-scale, diverse pathology datasets that span multiple institutions, staining variations, and scanner types^45^. Unlike general computer vision SSL models that benefit from extensive public datasets such as ImageNet ^20^ or LAION-5B ^46^, pathology-specific SSL models often lack sufficient pre-training diversity, making them susceptible to dataset-specific artifacts.

A systematic comparison is needed to evaluate pathology foundation models against both domain-specific SSL models and general computer vision SSL models, assessing their robustness across independent external datasets. Additionally, benchmarking should consider model generalization across diverse tissue types and cancer subtypes, ensuring that performance gains are not restricted to specific datasets. In this study, we compare general vision models (VM), general vision-language models (VLM), pathology-specific vision models (Path-VM), and pathology-specific vision-language models (Path-VLM), to analyze their generalization capability and task-specific strengths.

Altogether, over 20 pathology or pathology-language models have been released. In addition, several studies^47^ have used off-the-shelf imagenet models such as ResNet, DINO and DINOv2^38,39,48,49^ for medical imaging including digital pathology. However, a comprehensive comparison of these models across diverse datasets and tasks remains lacking. Here, we present such a benchmark of four different types of models: pathology image, vision (i.e. natural images), pathology-language and vision-language models to identify the best-performing foundation models on 41 downstream tasks, 53 datasets and over 17.5k samples. We conduct a comprehensive evaluation by testing them across identical downstream tasks within a standardized experimental framework, ensuring a fair comparison. Our analysis evaluates the consistency of top-performing models, identifies those excelling across diverse benchmarking tasks and tissue types, compares the efficacy of pathology image models against language and vision models, and examines the impact of model size and dataset size on performance. Finally, we explored the potential of fusion models to enhance generalization across external tasks and diverse tissues. This work aims to provide insights into the factors driving effective pathology foundation models, offering a foundation for optimizing their application in precision medicine.

## Results

### Benchmarking models on TCGA: Task-specific performance highlights variability in top model rankings

We conducted an extensive benchmarking study of 31 models across diverse domains, including general vision, general vision-language, pathology-specific vision, and pathology specific vision-language, evaluating them over 41 distinct tasks. To ensure consistent and concise terminology throughout the study, we assigned model abbreviations based on their training domains and modalities (Table 1): general vision models (VM), general vision-language models (VLM), pathology-specific vision models (Path-VM), and pathology-specific vision-language models (Path-VLM). Among these, 19 tasks were curated from the TCGA dataset, which includes multiple tumor types such as brain, breast, bladder, and others. We evaluated model performance using the average of balanced accuracy, precision, recall, and F1 score, collectively defined as the average performance metric for a comprehensive assessment. Initially, we focused on 15 pathology-specific vision models (Path-VM, Table 2), tested across 19 tasks related to tumor subtyping, molecular subtyping, and grading, and calculated the mean of average performance across 19 tasks.

**Table 1.** Abbreviations of Model Categories.

| Model Category | Model Category Abbreviation | Training data |
| --- | --- | --- |
| General vision models | VM | Natural images |
| General vision language models | VLM | Natural images and text |
| Pathology-specific vision models | Path-VM | Pathology-specific images |
| Pathology-specific vision language models | Path-VLM | Pathology-specific images and text |

**Table 2.**
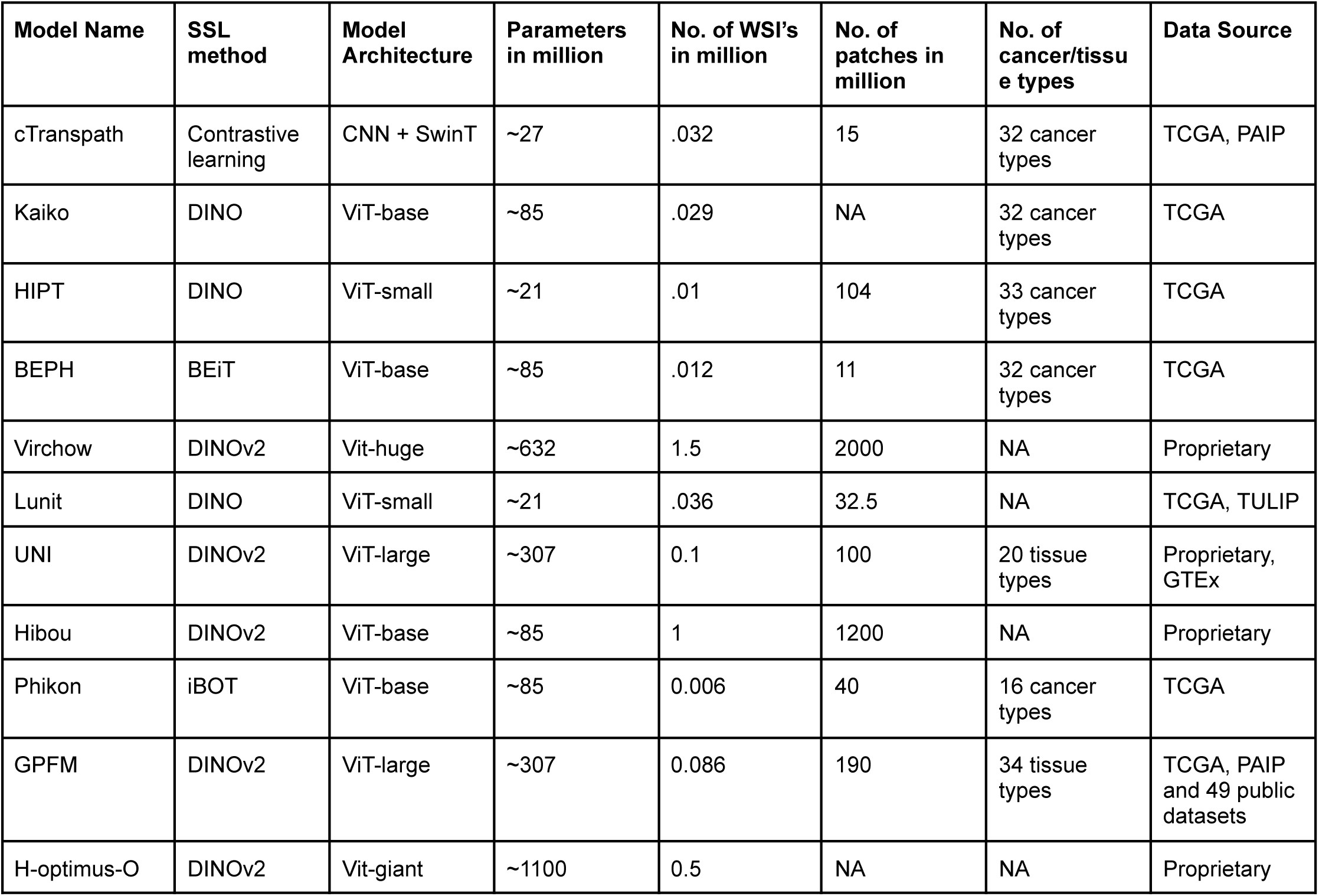
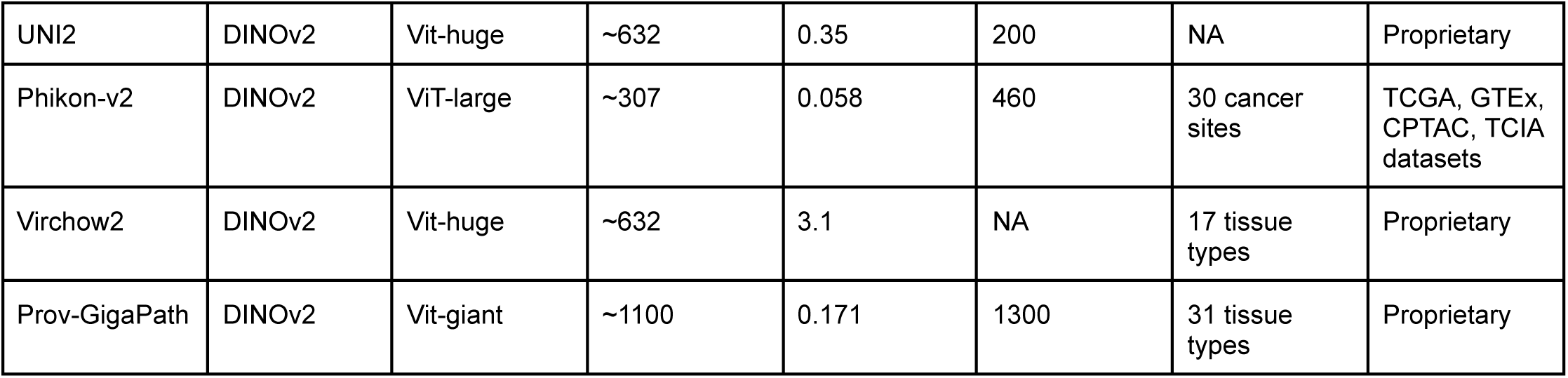
Pathology-specific vision model (Path-VM) summary.

**Table 3.** Pathology-specific vision-language model (Path-VLM) summary.

| Model Name | Pretraining method | Model Architecture | Parameters in million | Data source |
| --- | --- | --- | --- | --- |
| PLIP | CLIP | ViT-base | ~85 | 208k pathology image-text pairs |
| QuiltNet-B16 | CLIP | ViT-base | ~85 | 1M pathology image-text pairs |
| BiomedCLIP | CLIP | ViT-base | ~85 | 14M pathology image-text pairs |
| Mizero pubmedbert | Contrastive learning | ViT-small | ~21 | 33k pathology image-text pairs |
| Mizero clinmedbert | Contrastive learning | ViT-small | ~21 | 33k pathology image-text pairs |
| CONCH | CoCa<sup>77</sup> | ViT-base | ~85 | 1.17 pathology image-text pairs |
| TITAN | iBOT + CoCa<sup>77</sup> | ViT-base | ~85 | 336k whole slide images & 423k image captions |

**Table 4.** General Vision Model (VM) summary.

| Model Name | SSL method | Model Architecture | Parameters in million | Data source |
| --- | --- | --- | --- | --- |
| Dino-S16 | DINO | ViT-small | ~21 | ImageNet (14M) |
| Dino-B16 | DINO | ViT-base | ~85 | ImageNet (14M) |
| Dinov2 | DINOv2 | ViT-large | ~300 | LVD-142M |
| iBOT-B16 | iBOT | ViT-base | ~85 | ImageNet (14M) |
| iBOT-L16 | iBOT | ViT-large | ~307 | ImageNet (14M) |

**Table 5.** General Vision-language model (VLM) summary.

| Model Name | Pretraining method | Model Architecture | Parameters in million | Data source |
| --- | --- | --- | --- | --- |
| CLIP-B16 | CLIP (Contrastive learning) | ViT-base | ~115 | 400 million (image, text) pairs |
| BLIP-B16-14M | BLIP | ViT-base | ~85 | ImageNet (14M) |
| Align-base | EfficientNet + BERT (Contrastive learning) | ViT-base | ~746 | 700 million (image, text) pairs |
| beit3-L16 | iBOT | ViT-large | ~307 | ImageNet (14M) |

**Table 6.** Downstream tasks.

|  | Task | Cancer | Binary or Multi | Dataset | Slide or patch classification | tissue |
| --- | --- | --- | --- | --- | --- | --- |
| 1 | TCGA-Grade | Pan-cancer | multi-class | TCGA | Slide | pan-cancer |
| 2 | PAM50 status | BRCA | multi-class | TCGA | Slide | breast |
| 3 | prad mrna cluster | PRAD | multi-class | TCGA | Slide | prostate |
| 4 | IDH mutation | GBM + LGG | binary | TCGA | Slide | brain |
| 5 | brain hist subtype | GBM + LGG | multi-class | TCGA | Slide | brain |
| 6 | PI3K | Pan-cancer | binary | TCGA | Slide | pan-cancer |
| 7 | ER status | BRCA | binary | TCGA | Slide | breast |
| 8 | MYC | Pan-cancer | binary | TCGA | Slide | pan-cancer |
| 9 | Immunegroup | Pan-cancer | multi-class | TCGA | Slide | pan-cancer |
| 10 | MGMT | GBM + LGG | binary | TCGA | Slide | brain |
| 11 | blca hist subtype | BLCA | multi-class | TCGA | Slide | brain |
| 12 | gi subtype | GI | multi-class | TCGA | Slide | GI |
| 13 | brain transcriptome subtype | GBM + LGG | multi-class | TCGA | Slide | brain |
| 14 | lung transcriptome subtype | LUSC + LUAD | multi-class | TCGA | Slide | lung |
| 15 | blca mrna cluster | BLCA | multi-class | TCGA | Slide | bladder |
| 16 | lusc vs luad | LUSC + LUAD | binary | TCGA | Slide | lung |
| 17 | ERG status | PRAD | binary | TCGA | Slide | prostate |
| 18 | kidney subtype | KIRC + KIRP + KICH | multi-class | TCGA | Slide | kidney |
| 19 | PR status | BRCA | binary | TCGA | Slide | breast |
| 20 | CPTAC-grade | Pan-cancer | multi-class | CPTAC | Slide | pan-cancer |
| 21 | SWI | Pan-cancer | binary | CPTAC | Slide | pan-cancer |
| 22 | P53 | Pan-cancer | binary | CPTAC | Slide | pan-cancer |
| 23 | Chrom | Pan-cancer | binary | CPTAC | Slide | pan-cancer |
| 24 | MTOR | Pan-cancer | binary | CPTAC | Slide | pan-cancer |
| 25 | MYC | Pan-cancer | binary | CPTAC | Slide | pan-cancer |
| 26 | tumor label | Pan-cancer | multi-class | CPTAC | Slide | pan-cancer |
| 27 | SICAPv2 | PRAD | multi-class | External benchmarking | Patch | prostate |
| 28 | MHIST | COAD | binary | External benchmarking | Patch | colon |
| 29 | UniToPatho | COAD | multi-class | External benchmarking | Patch | colon |
| 30 | BACH | BRCA | multi-class | External benchmarking | Patch | breast |
| 31 | BRACS | BRCA | multi-class | External benchmarking | Patch | breast |
| 32 | LC25000 | Pan-cancer | multi-class | External benchmarking | Patch | pan-cancer |
| 33 | BreakHis | BRCA | multi-class | External benchmarking | Patch | breast |
| 34 | NCT-CRC-HE | COAD | multi-class | External benchmarking | Patch | colon |
| 35 | lung stage | Lung | multi-class | Out of Domain (NLST) | Slide | lung |
| 36 | lung grade | Lung | multi-class | Out of Domain (NLST) | Slide | lung |
| 37 | lung hist OOD1 | Lung | binary | Out of Domain (Stanford) | Slide | lung |
| 38 | lung hist OOD2 | Lung | multi-class | Out of Domain (DHMC) | Slide | lung |
| 39 | Lung stage OOD | Lung | multi-class | Out of Domain (Stanford) | Slide | lung |
| 40 | Brain necrosis | Brain | binary | Out of Domain (Stanford) | Slide | brain |
| 41 | MGMT OOD | Brain | binary | Out of Domain (Stanford) | Slide | brain |

Among these, the Virchow2^28^ model consistently ranked first, achieving a mean average performance of 0.706 ± 0.10. The next best-performing models were H-optimus-0 (0.702 ± 0.11), Prov-GigaPath^50^ (0.702 ± 0.10) and UNI (0.70 ± 0.10), comprising the top four rankings (Figure 1b). Among the Path-VM, Virchow2 and UNI2 ranked first in 5 out of 19 TCGA-based tasks (Figure 1c), while H-optimus-0 and Prov-GigaPath ranked first in 3/19 tasks.

**Figure 1.**
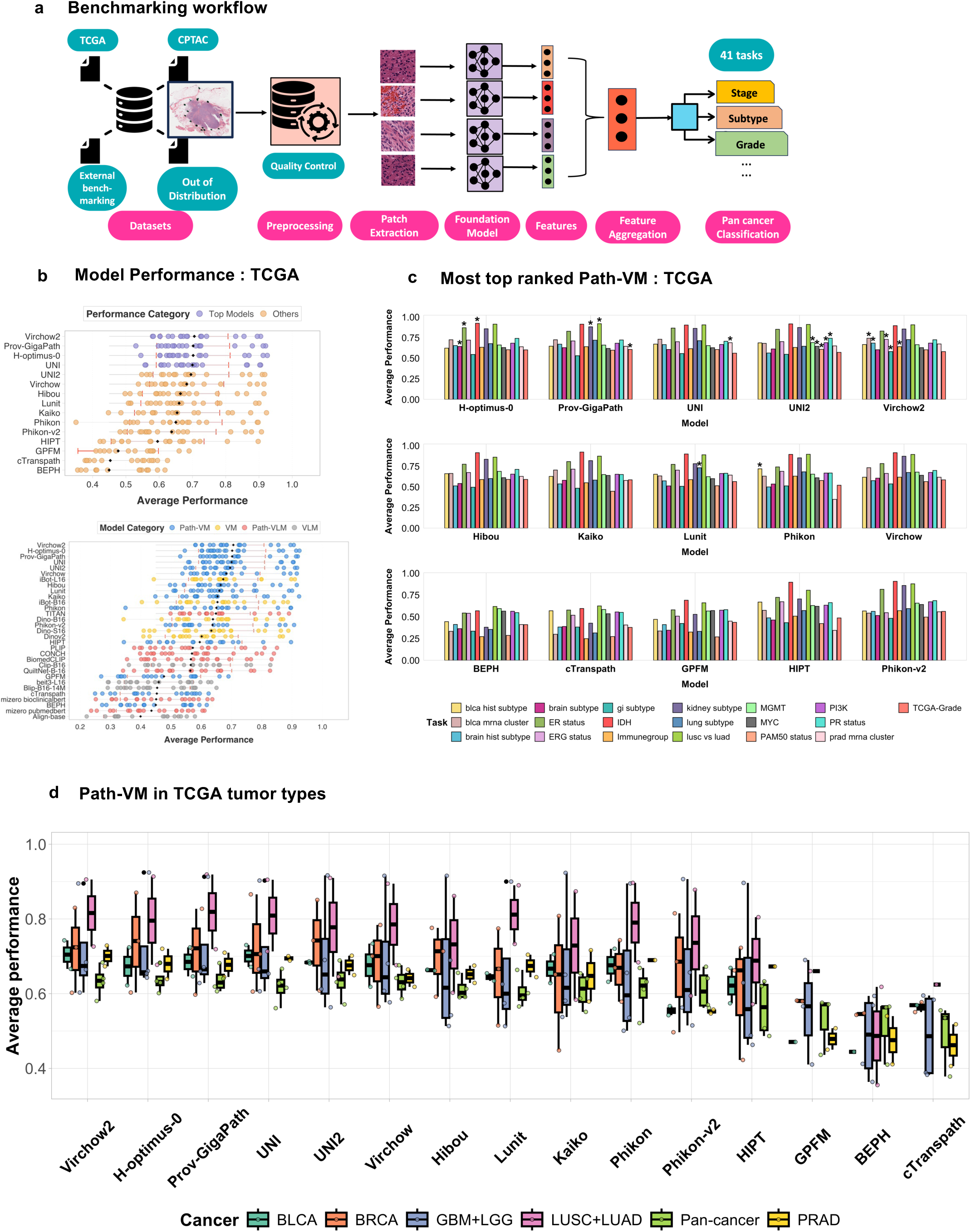
**a)**. **Benchmarking Workflow** A schematic representation of the benchmarking workflow used in this study. The framework includes data preprocessing, model feature extraction, evaluation on multiple histopathology datasets, and performance comparison across tumor classification, molecular subtyping, and grading tasks. **b). Model Performance on TCGA Dataset** Performance comparison of foundation models on TCGA-derived tasks. The top panel displays the average performance of Path-VM models, while the bottom panel includes all evaluated models including Path-VM, VM, VLM, Path-VLM. Average performance is defined as the mean of balanced accuracy, F1-score, recall, and precision. Each black dot indicates the mean performance across TCGA-derived tasks for a given model, while the red bars represent the standard deviation across those tasks. **c). Top-Ranked Path-VM Models** The most top ranked Path-VM model based on average performance metrics (marked with asterisk), demonstrating superior predictive performance across tumor classification, molecular subtyping, and grading tasks. The figure illustrates the frequency with which each model achieved the top ranking across 19 TCGA tasks. Each bar represents a specific TCGA task. **d)**. **Path-VM Performance Across Tumor Types** Evaluation of pathology-specific vision models across different tumor types, illustrating variations in predictive power depending on tissue origin and diagnostic complexity. Performance trends highlight strengths and limitations in pathology model generalization across tumor subtypes.

We further investigated whether specific models excel in tumor-type-specific classification and subtyping tasks, aiming to identify tailored strengths across diverse cancer contexts. H-optimus-0 demonstrated superior performance on brain and breast tumor classification tasks (Figure 1d, Supplementary Figure 1a,b). In contrast, Virchow2 outperformed others on bladder and prostate cancer subtyping tasks, showcasing its precision in delineating subtle histological subtypes critical for these cancers (Supplementary Figure 1b). Meanwhile, Prov-GigaPath achieved the highest rankings on lung cancer and pan-cancer assessments, excelling on both organ-specific and cross-cancer evaluations (Supplementary Figure 1a, b).

Notably, UNI (0.70 ± 0.10) outperformed UNI2 (0.69 ± 0.11) when evaluated by the mean average performance metric across all 19 tasks(Figure 1b), reflecting its consistent strength across diverse challenges. However, UNI2 secured the top rank on 5 individual tasks (Figure 1c), surpassing UNI, which achieved the top rank on only 1 task, highlighting a striking contrast in task-specific excellence versus overall consistency across the 19-task evaluation. A detailed ranking of the top 10 Path-VM across all TCGA tasks is provided in Supplementary Figure 2a, offering a comprehensive perspective on model performance. Next, when broadening the evaluation to include all models—spanning general vision (VM), general vision-language (VLM), pathology-specific vision models (Path-VM), and pathology-specific vision language (Path-VLM) architectures—the top 10 rankings remained dominated by Path-VM. The only exception was iBOT-L16^23^, a vision model, which emerged as the sole non-pathology model in this group (Figure 1b, bottom panel). Overall, our results show significant variability in top model rankings across tumor-type classification and subtyping challenges. We also developed a live benchmarking dashboard PathBench, accessible at [xx], to enable real-time visualization and comparison of model performance.

### Virchow2 is the most consistent pathology model in CPTAC and external benchmarking tasks

Next, we evaluated these foundation model categories (VM, Path-VM, VLM, Path-VLM) on CPTAC cancer cases spanning various tumor types, focusing on specific tasks such as tumor grading, tumor classification, and pathway activity prediction (e.g., p53, MTOR, SWI). Among the 31 models tested, Virchow2 demonstrated the highest overall performance (Figure 2a, Supplementary Figure 3c), followed closely by Prov-GigaPath and UNI, when averaged across seven CPTAC-based tasks. Virchow2 achieved the best performance in 2 out of 6 individual tasks (Supplementary Figure 3a), with an average score of 0.766 ± 0.103, while Prov-GigaPath and UNI followed with 0.763 ± 0.103 and 0.760 ± 0.103, respectively. Notably, Prov-GigaPath also excelled in 2 out of the 6 tasks, with an average performance of 0.763 ± 0.11. Both Virchow2 and Prov-GigaPath consistently ranked among the top three models across CPTAC tasks (Supplementary Figure 3a). H-optimus-0, the second-best model in TCGA-based tasks, ranked fourth in CPTAC tasks. In the tumor type classification task, Prov-GigaPath achieved the highest performance, followed by Virchow and Virchow2.

**Figure 2.**
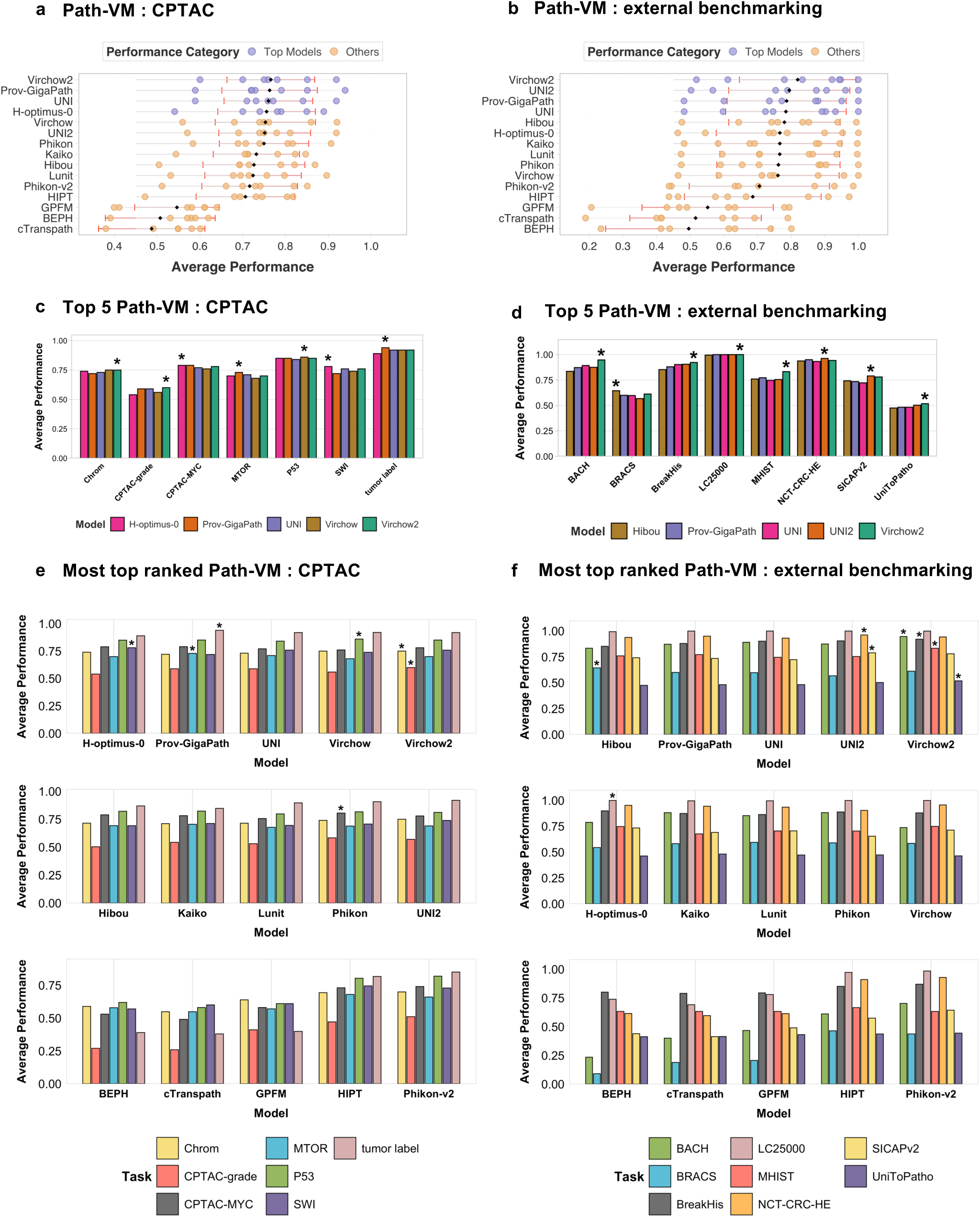
**a). Path-VM performance in CPTAC** – Performance comparison of pathology-specific vision models on CPTAC datasets. Models are evaluated based on average performance across multiple classification tasks, highlighting their robustness and generalizability within the CPTAC domain. **b). Path-VM performance in External benchmarking** – Evaluation of pathology models on external benchmarking datasets, assessing their ability to generalize beyond training distributions and perform effectively on independent datasets. **c). Top 5 Path-VM performance in CPTAC** – Performance analysis of the five highest-ranking Path-VM on CPTAC datasets, demonstrating their relative strengths across classification tasks. The top performing model for a task is highlighted with an asterisk. **d). Top 5 Path-VM Performance in External Benchmarking** – Comparative performance of the top five Path-VM on external benchmarking datasets, illustrating their generalizability across diverse histopathology data sources. The top performing model for a task is highlighted with an asterisk. **e). Most top-ranked Path-VM in CPTAC** among all Path-VM– The number of times each Path-VM achieved the highest ranking across CPTAC classification tasks. Each bar represents a specific CPTAC task. The asterisk represents the top performing model in that task. **f). Most top-ranked Path-VM in External benchmarking** – The frequency of top rankings achieved by Path-VM across external benchmarking tasks. Each bar represents a distinct classification task, highlighting the best-performing models in out-of-domain evaluations.

Next, we evaluated these model categories (VM, Path-VM, VLM, Path-VLM) across eight external benchmarking tasks spanning multiple cancer types, including breast, prostate, and colorectal cancer. Virchow2 outperformed all other models, achieving an average performance of 0.82 ± 0.17 (Figure 2b, Supplementary Figure 3d), followed by UNI2 (0.79 ± 0.18). Prov-GigaPath ranked third, with a mean performance of 0.787 ± 0.17 across these tasks. Notably, Virchow2 consistently ranked among the top three in 6 out of 8 external benchmarking tasks. In contrast, H-optimus-0, which ranked second in TCGA-based tasks based on mean performance, performed poorly in external benchmarking tasks, placing seventh (0.766 ± 0.18). Interestingly, UNI2 outperformed its counterpart UNI in external tasks (Supplementary Figure 3d), where UNI ranked fourth (0.784 ± 0.18). However, in CPTAC tasks, UNI (0.760 ± 0.10, ranked third) outperformed UNI2 (0.751 ± 0.10, ranked sixth). We assessed whether the top five models from TCGA-based tasks demonstrated statistically significant performance differences compared to the remaining models across the three dataset categories. Virchow2 was the only model to show statistically significant performance gains in both CPTAC and external benchmarking tasks (Supplementary Figure 4a). UNI also exhibited statistically significant differences in CPTAC tasks; however, it did not show significant improvement in external benchmarking tasks (Supplementary Figure 4b). Overall, Virchow2 demonstrated consistently strong performance across both CPTAC and external benchmarking tasks.

### Pathology-specific vision models (Path-VM) achieve superior performance over pathology-specific vision language models (Path-VLM) and general vision models (VM)

Next, we systematically evaluated the performance of pathology-specific vision models (Path-VM), pathology-specific vision-language models (Path-VLM), and general vision models (VM) across classification tasks from TCGA, CPTAC, and eight external benchmarking datasets (Figure 3a). In comparative analysis of Path-VM and VM, no statistically significant differences were observed between Path-VM and VM across these datasets (Figure 3a). To further assess their practical effectiveness, we analyzed model rankings, where Path-VM’s superior visual feature processing translated into consistent dominance across tasks (Figure 3b,c). Path-VM consistently dominated the top 10 positions in combined comparisons of VM and Path-VM, underscoring their practical superiority (Figure 3b,c, Supplementary Figure 8a, Supplementary Figure 9a,b). In 19 TCGA tasks, Path-VM held nearly all top 10 positions, with only a single VM appearing in most cases (Supplementary Figure 8a, Supplementary Figure 10b). Similarly, in CPTAC and external tasks (Figure 3b,c), Path-VM were prevalent among the top 10, with only a few VM achieving high rankings (Supplementary Figure 8a-b). For example, iBOT VM achieved the second-highest performance in the TCGA brain histology subtype classification task and TCGA gastrointestinal subtype classification (Supplementary Figure 8a). A few VM’s(iBOT and DINO based general vision models) also ranked among the top three in the colorectal cancer challenge (MHIST), and the UnitoPatho challenge, and the DINO-B16 VM ranked fourth in the CPTAC-chromatin modification pathway classification task associated with gastrointestinal and colorectal tumors (Supplementary Figures 9a-b). These isolated successes suggest that VM’s can learn meaningful features for specific tasks, such as colorectal and gastrointestinal tumors, but Path-VM’s consistent top rankings highlight their broader superiority.

**Figure 3.**
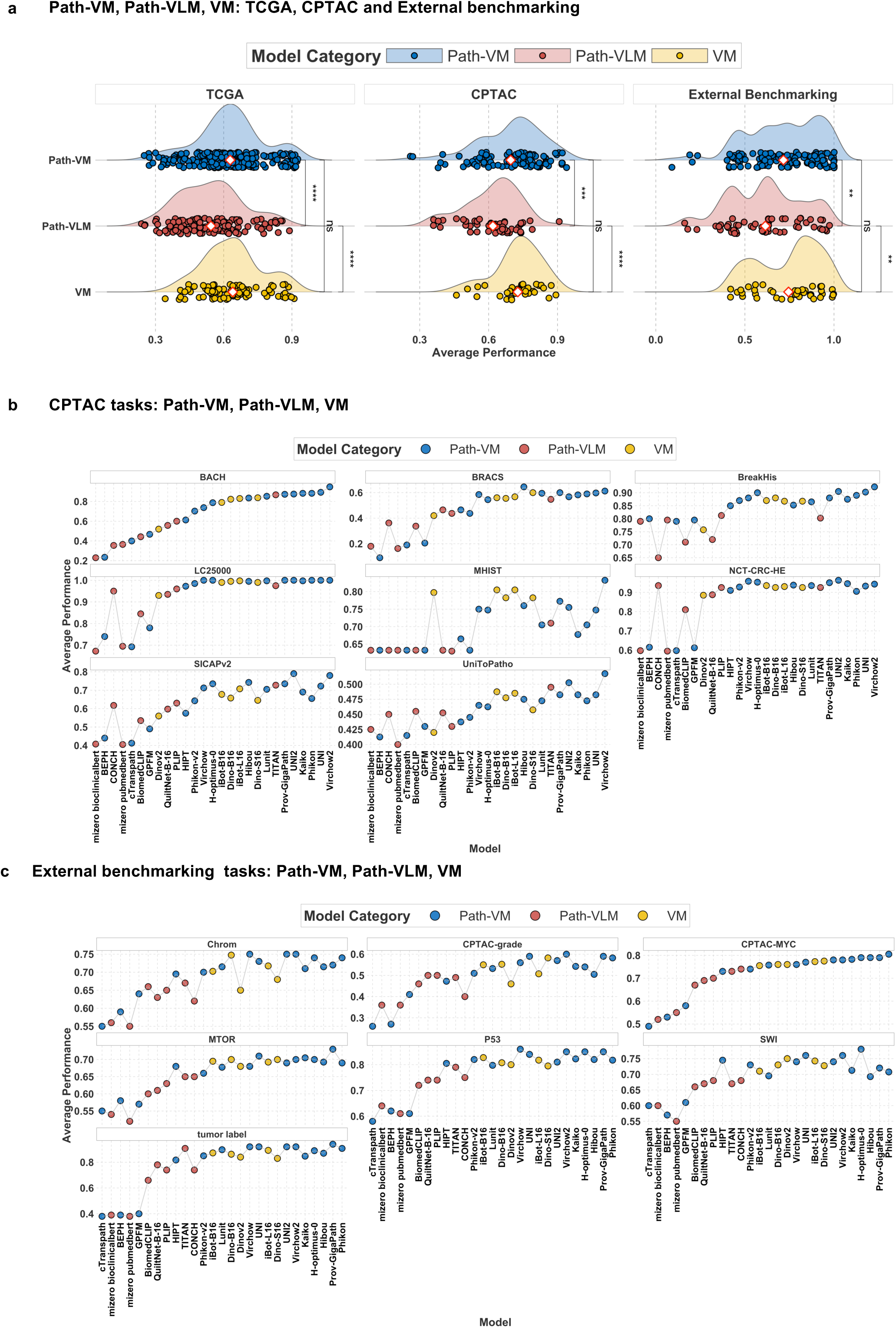
**a). Path-VM, Path-VLM and VM in TCGA.** Comparison of average performance between pathology-specific vision models (Path-VM), pathology-specific vision-language models (Path-VLM) and general vision models (VM) across TCGA, CPTAC and External benchmarking tasks. Path-VM perform better than Path-VLM across TCGA, CPTAC and External benchmarking tasks, with statistically significant differences. No statistically significant differences observed between Path-VM and VM. **b). Path-VM, Path-VLM and VM in each CPTAC task.** Per-task performance of Path-VM, Path-VLM and VM across CPTAC datasets. Each dot represents a model’s performance in a given task, illustrating the consistent advantage of pathology-specific vision models across most tasks. **c)**. **Path-VM, Path-VLM and VM in each External benchmarking task.** Per-task performance comparison between Path-VM, Path-VLM and VM on external datasets. Path-VM consistently outperform and dominate top rankings, demonstrating superior performance.

In a comparative analysis of Path-VM and Path-VLM models across pathology tasks, Path-VM consistently outperformed Path-VLM in TCGA, CPTAC, and external benchmarking datasets, with statistical significance (Figure 3a). In TCGA tasks, only a few Path-VLM were ranked in top 10 in the combined ranking of Path-VM and Path-VLM (Supplementary Figure 5a, Supplementary Figure 10a) such as BiomedCLIP and QuiltNet-B-16 in bladder histology subtyping and MGMT methylation prediction, TITAN in brain histology, brain subtype(transcriptomic) and gi subtyping tasks. Similarly, in CPTAC tasks, Path-VLM failed to consistently rank within the top 10. The only exception was TITAN, a Path-VLM, which achieved a 6th-place ranking in the tumor label classification task (Supplementary Figure 6a). In external benchmarking tasks, Path-VLM again showed limited success. The only exception,TITAN, did achieve top-10 rankings in five out of eight tasks and ranked among the top three in the UnitoPatho challenge (Supplementary Figure 7a).

A comparative analysis of vision-only models (VM and Path-VM), and vision-language models (VLM and Path-VLM), revealed that vision-only models, both VM and Path-VM significantly outperformed VLM and Path-VLM across TCGA, CPTAC, and external benchmarking tasks (Supplementary Figures 11a-c). While Path-VLM exhibited a statistically significant performance advantage over VLM due to pathology-specific textual data, both vision-language models lagged behind their vision-only counterparts. Even though VLMs are trained on large datasets, they might have underperformed due to diluted visual feature learning from multimodal objectives and insufficient pathology-specific pretraining. This highlights the pivotal role of high-resolution visual features in pathology image analysis, where models relying solely on image data consistently excel over those incorporating textual information.

Overall, these results highlight the practical advantages of pathology-specific vision models, which leverage robust visual representations of tissue morphology to achieve superior classification performance, as demonstrated by Path-VM’s statistically significant outperformance of Path-VLM. Path-VM’s dominance in top 10 rankings, even in the absence of statistical significance over VM, underscores the value of pathology-specific training on larger, more diverse datasets that better represent the target tasks. While VM demonstrate potential in specific tasks, their limited presence in top rankings compared to Path-VM highlights the advantage of specialized models. Path-VLM, trained on histopathology slides and textual data (For example, CONCH^36^ trained on 1.17 million image-text pairs, and PLIP^34^ on ∼200k image-text pairs), generally underperformed, likely due to smaller, less diverse training datasets that may not fully leverage the complementary nature of image and text modalities for classification tasks. The underperformance of Path-VLM may result from limitations in their current architectures or training data, which may hinder their effectiveness in classification tasks, suggesting a need for improved multimodal integration to achieve performance comparable to Path-VM.

### Model size and data size do not guarantee better performance in pathology foundation models

We first investigated model size and specifically whether larger vision transformer (ViT) architecture (i.e. ViT-large, ViT-huge, and ViT-giant collectively referred to as ViT-L and model parameters > 300M) offer superior performance compared to small(model parameters = 21M) and base ViT (model parameters = 85M) among pathology models. In TCGA-based tasks, larger ViT architectures outperformed both ViT-S and ViT-B models (Figure 4a). However, for CPTAC and external benchmarking datasets, no statistically significant performance differences were observed among the different ViT sizes (Figure 4a). Similarly, when evaluating vision models, we found no statistically significant differences in performance among ViT-S, ViT-B, and ViT-L across TCGA, CPTAC, and external benchmarking tasks (Supplementary Figures 12a-c).

**Figure 4.**
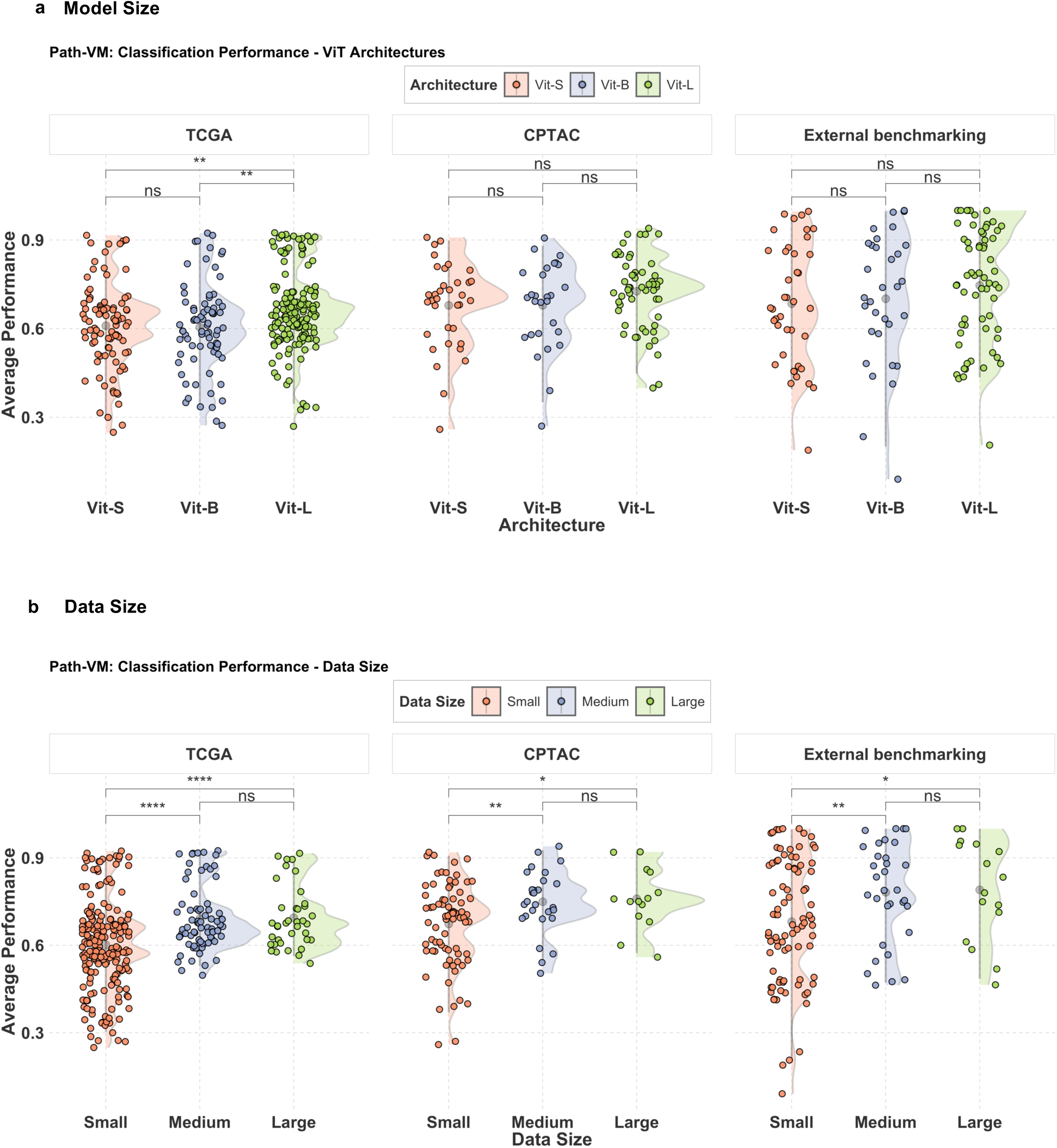
**a) Path-VM : Comparison of ViT Architectures in TCGA, CPTAC and External benchmarking** – Model performance analysis of ViT architectures on TCGA tasks reveals statistically significant differences between ViT-small, and ViT-large models, as well as between ViT-base and ViT-large models, while no significant difference is observed between ViT-small, and ViT-base models. Model performance evaluation on CPTAC tasks indicates no statistically significant differences among ViT-small, ViT-base, and ViT-large architectures. Performance comparison of ViT architectures across external benchmarking datasets shows no statistically significant differences among ViT-small, ViT-base, and ViT-large models. **b) Path-VM**: **Impact of pretraining data Size on performance in TCGA, CPTAC and External benchmarking** – Model performance in TCGA classification tasks as a function of the number of whole-slide images used during pretraining, highlighting the influence of data scale on downstream accuracy. Data sizes are categorized as small (<0.1 million slides), medium (0.1 - 1 million slides), and large (≥1 million slides). Statistical analysis reveals significant differences between small and large as well as small and medium data sizes, while no significant difference is observed between medium and large data sizes. Statistical analysis of model performance in CPTAC tasks reveals significant differences between small and large data sizes, as well as between small and medium, but no significant difference between medium and large. Statistical analysis of model performance in External benchmarking indicates significant differences between small and large data sizes, as well as between small and medium, with no significant difference observed between medium and large data sizes in the external benchmarking tasks. Significance levels: * p < 0.05, ** p < 0.01, *** p < 0.001 (pairwise t-tests with Bonferroni correction)

To assess the impact of pre-training dataset size on model performance, we categorized pathology models based on the number of whole-slide images (WSIs) used for training: small (<0.1 million WSIs), medium (0.1–1 million WSIs), and large (>1 million WSIs). Our analysis revealed statistically significant differences in average performance between models trained on small versus medium datasets, as well as small versus large datasets, across TCGA, CPTAC, and external benchmarking tasks (Figure 4b). However, no statistically significant difference was observed between models trained on medium and large datasets (Figure 4b). For example, models such as UNI (trained on ∼100k WSIs) and UNI2 (∼300k WSIs) achieved top performance across multiple tasks, were close behind the performance of Virchow2, which was trained on 3 million WSIs, (Figure 1b,c, Figure 2a,b). Additionally, UNI, trained on ∼100k WSIs, and H-optimus-0, trained on ∼500k WSIs, outperformed both Hibou and Virchow, which were trained on ∼1 million WSIs and 1.5 million WSI’s respectively, across all three dataset categories (Figure 1b,c and Figure 2a,b). These findings suggest that beyond a certain threshold, increasing training data size does not necessarily lead to improved classification performance and that diversity in the training set may play a bigger role in models’ performance.

### Fusion model generalizes better across external tasks and tissues

Next, we evaluated model performance using a combination of proprietary and out-of-domain datasets (i.e. Stanford, NLST, and DHMC) as part of an out-of-domain assessment, alongside external tasks to assess generalizability. Since these datasets were not included in model training, they served as an external evaluation of model robustness. We ranked the models based on their cumulative average performance across TCGA, CPTAC, and a combined set of out-of-domain and external benchmarking tasks. While H-Optimus-0 (0.702 ± 0.11) and Prov-GigaPath (0.702 ± 0.10) ranked among the top-performing models in 19 TCGA tasks (Figure 5a), their average performance showed a moderate decline in out-of-domain and external benchmarking tasks, with scores of 0.662 ± 0.19 and 0.662 ± 0.21, respectively (Figure 5b), leading to lower rankings. Similarly, UNI2 (0.696 ± 0.11) and Virchow (0.683 ± 0.11) exhibited a slight decrease in average performance from TCGA tasks (Figure 5a) to 0.664 ± 0.21 and 0.662 ± 0.20 in external evaluations (Figure 5b). These shifts in rankings across datasets highlight variability in model generalizability. To explore whether combining top-performing foundation models could improve predictive performance, we applied a late fusion strategy that aggregates predictions using majority voting. We selected the top five models based on mean average performance across TCGA tasks (Figure 1b): Virchow2, H-Optimus-0, Prov-GigaPath, UNI, and UNI2. We then compared the fusion model’s performance against individual models across multiple evaluation categories, including TCGA, CPTAC, and external benchmarking datasets (which included out-of-domain tasks). In TCGA tasks, the best-performing individual model, Virchow2, surpassed the fusion model in cumulative average performance across 19 tasks (Figure 5a). However, in CPTAC and external evaluations, the fusion model outperformed all individual models (Figures 5b,c), suggesting stronger generalizability.

**Figure 5.**
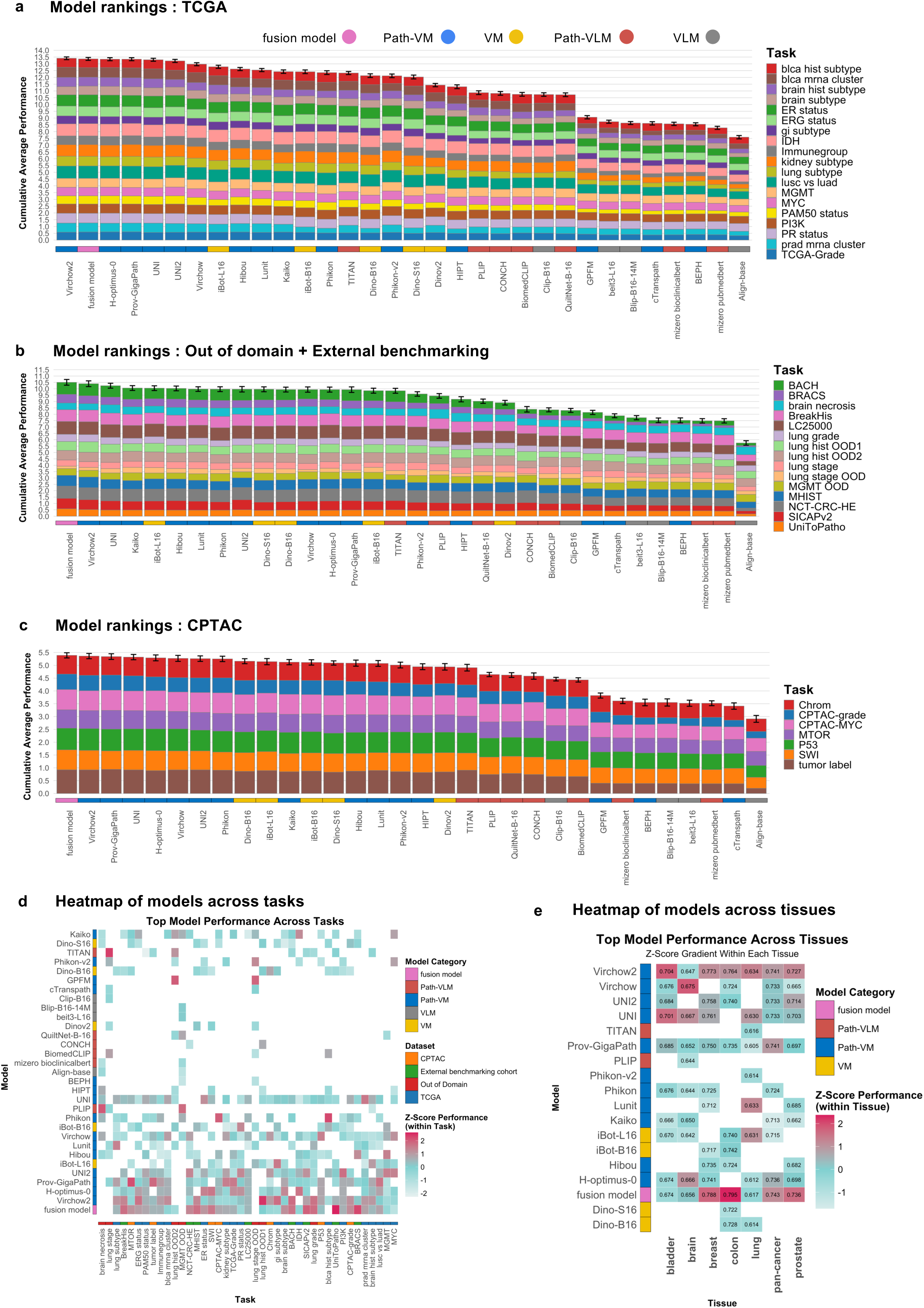
**a) Model Rankings: TCGA** – Cumulative average performance of pathology models across TCGA classification tasks, with rankings reflecting overall model performance. The fusion model ranks second in this category, demonstrating strong performance across TCGA tasks. **b) Model Rankings: External Benchmarking + Out-of-Domain** – Cumulative average performance of pathology models across external benchmarking and out-of-domain classification tasks. This ranking emphasizes the ability of models to generalize across datasets that were not used during pre-training, with the fusion model achieving the top ranking in this category. **c) Model Rankings: CPTAC** – Cumulative average performance of pathology models across CPTAC classification tasks, showcasing the overall model rankings within the CPTAC dataset. The fusion model performs the best in this category, leading the rankings across CPTAC tasks. **d) Heatmap of Model Performance Across All Tasks** – Heatmap showing model performance across all classification tasks, with z-scores indicating performance relative to other models within each specific task. This provides insight into how models perform across a broad range of pathological challenges. **e) Heatmap of Model Performance Across Tissues** – Heatmap displaying model performance across various tissue types, with z-scores calculated for each model’s performance within individual tissue categories. This illustrates how each model performs relative to other models for each tissue type.

To further analyze model specialization, we calculated z-scores for each model’s performance and identified the top 10 models per task. Grouping all 41 tasks by tissue type (Figure 5d), we found that the fusion model excelled in breast, colon, prostate, and pan-cancer tasks, while Virchow2 performed best in bladder and lung cancer tasks. A direct comparison of Virchow2 and the fusion model (Figure 5e) showed that in TCGA tasks, Virchow2 outperformed the fusion model in 11 of 19 tasks. However, in CPTAC and external evaluations, the fusion model ranked higher in 5 of 7 CPTAC tasks and 12 of 15 out-of-domain + external tasks, suggesting its ability to generalize across diverse datasets. The fusion model exhibited superior generalizability, outperforming individual models in CPTAC and external tasks, particularly in breast, colon, prostate, and pan-cancer classifications. It achieved the highest overall performance across datasets, emphasizing the strength of ensemble-based predictions.

## Discussion

This study presents an in-depth benchmarking analysis of 31 models spanning multiple domains—general vision models (VM), general vision-language models (VLM), pathology-specific vision models (Path-VM), and pathology-specific vision-language models (Path-VLM)—evaluated across 41 distinct tasks, which include a comprehensive range of cancer classification and subtyping challenges derived from TCGA, CPTAC, and external benchmarking datasets. The goal of this study was to investigate the task-specific strengths of various models and assess their overall performance on common cancer-specific tasks. Our findings reveal notable variations in model performance, reflecting the inherent complexity of pathology tasks and emphasizing the necessity for tailored approaches depending on the type of cancer and the specific challenge. In our study, it is crucial to also acknowledge that TCGA (The Cancer Genome Atlas)^30^ likely serves as a major source of training data for many of the models evaluated. Despite the use of self-supervised learning (SSL) techniques aimed at minimizing reliance on labeled data, some degree of overfitting on TCGA-specific characteristics may still occur. This is particularly relevant when evaluating models on TCGA-based tasks, as the overlap between training and evaluation datasets can lead to inflated performance results, making it essential to consider this factor when interpreting the findings.

Virchow2 and UNI emerged as the top-performing pathology-specific vision models (Path-VM) across TCGA, CPTAC, and external tasks, with Virchow2 demonstrating the highest consistency and excelling in distinguishing subtle histological subtypes critical for cancers such as bladder and prostate. Meanwhile, H-optimus-0 and Prov-GigaPath exhibited strong performance in specific tumor types—H-optimus-0 excelled in brain and breast cancer classification, while Prov-GigaPath was highly effective in lung cancer and pan-cancer evaluations. A key observation from this study is the variability in model performance across out-of-domain and external tasks; while models such as H-optimus-0, Prov-GigaPath, and UNI2 performed well on TCGA tasks, their performance declined on external datasets, underscoring the challenge of ensuring generalizability beyond the training dataset. This task-specific specialization suggests that models optimized for particular cancers may exhibit strong performance in niche domains, even if their overall rankings do not fully capture these targeted strengths. Fusion models proved particularly valuable in addressing these generalizability challenges—by aggregating predictions from top-performing individual models using majority voting, they demonstrated superior performance across external datasets, including CPTAC, even though they did not surpass the best individual model (Virchow2) on TCGA tasks. Their strong results in breast, colon, prostate, and pan-cancer classifications further highlight their ability to generalize across diverse tissue types and datasets, making them a promising avenue for enhancing model robustness and real-world applicability. Given that ensemble methods are often more resilient to unseen data^51–53^, fusion models offer a compelling strategy for improving diagnostic AI in pathology.

The comparatively lower performance of pathology-specific vision language models (Path-VLM) likely stems from their reliance on smaller datasets, as fewer sources contain both pathology images and paired textual annotations. Even though VLMs (general vision-language models) are trained on large datasets, such as those with hundreds of millions of image-text pairs, they underperform due to the mismatch between general-domain pretraining data and pathology-specific high-resolution images, noisy or irrelevant textual annotations, and limitations in handling the high-resolution visual details critical for pathology tasks. While these models integrate visual and linguistic information, their current architectures may not fully harness the complementary nature of these modalities, limiting their effectiveness in classification tasks. Advancements in multimodal learning, particularly with larger, well-curated datasets and improved fusion techniques, could enhance their capabilities and make them more competitive with image-specific models. Meanwhile, general vision-only models (VM) trained on large-scale datasets like ImageNet^20^ exhibited competitive performance in certain tasks but were generally outperformed by pathology-specific vision models (Path-VM) across TCGA, CPTAC, and external benchmarks. This highlights the unique domain-specific features of pathology vision data that general-purpose vision models, trained on non-medical images, fail to fully capture. While VM excelled in specific cancer subtypes, such as gastrointestinal and colorectal tumors, Path-VM demonstrated a more consistent advantage across a broader range of tasks, underscoring the importance of in-domain pre-training for developing more specialized and effective computational pathology models.

This study challenges the assumption that larger models and more training data always lead to better performance. While larger vision transformer architectures showed advantages in TCGA tasks, their gains did not extend to CPTAC and external datasets. Similarly, increasing the number of whole-slide images beyond a certain threshold did not consistently improve classification accuracy. Notably, smaller models performed competitively with or even outperformed some models trained on much larger datasets. These findings highlight that beyond a certain scale, model architecture, training strategies, and data quality are more critical than sheer size.

Overall, these findings provide valuable insights into the strengths of various model architectures and strategies for pathology-related tasks. The results highlight that, while generalization is a key challenge, task-specific model optimization, along with ensemble-based approaches, can significantly enhance model performance. Future work could focus on exploring the considerable potential of multimodal models that integrate both image and textual data, alongside strategies such as ensemble approaches, addressing data diversity^16,54^ and optimization techniques for pathology vision models (Path-VM), to enhance model transferability across diverse datasets.

## Methods

### Datasets for downstream tasks

#### TCGA dataset

The Cancer Genome Atlas (TCGA) project aimed at cataloging and generating comprehensive molecular landscapes responsible for cancer. The Cancer Genome Atlas project^30^ generated multiple omics modalities, including exome-seq, RNA-seq, and methylation data, along with digital pathology slides, covering over 32 tumor types and 10,000 patients. With respect to digital pathology, TCGA provided a vast collection of high-resolution whole-slide images of tumor sections stained with Hematoxylin and Eosin (H&E). This has facilitated the development of machine learning and deep learning algorithms for tumor subtype classification, prognostication, and treatment response prediction using pathology imaging data. We used this rich resource of data for evaluation of foundation models. We evaluated the models based on tumor-specific molecular subtyping, such as distinguishing between IDH mutant and wild-type brain tumors, as well as pan-cancer molecular subtyping, like differentiating between TP53 mutant and wild-type tumors. This approach provided an estimate of the models’ performance in specific tumors and across a pan-cancer setting. Additionally, we evaluated these models on a diverse range of tasks, including pathway activity, immune group subtyping, mRNA cluster classification, and histological subtyping, across various cancer types. Overall, we curated around 19 downstream tasks from TCGA data, encompassing bladder, brain, prostate tumors, and more.

#### CPTAC + External benchmarking datasets

The Clinical Proteomic Tumor Analysis Consortium ^55^(CPTAC) has provided pan-cancer datasets that include omics and digital pathology data, similar to the TCGA project. We also used this resource to evaluate our models. Since most pathology models have been trained on the TCGA dataset, testing their performance on CPTAC pathology data will offer a true measure of their efficacy on an independent test dataset. From CPTAC, we curated 7 pan-cancer tasks^56^ : grade, SWI pathway alteration, p53, chromatin modification pathway alteration, mTOR pathway alteration, MYC pathway alteration, tumor label. In addition to CPTAC, we curated 8 publicly available external benchmark datasets, including BRACS^57^, BACH^58^, UnitoPatho ^59^, SICAPv2^60^, BreakHis^61^, LC2500^62^, MHIST^63^, and NCT-CRC-HE^64^. These datasets contain representative images from diverse tumor types and are completely independent of the training data, ensuring a robust and unbiased evaluation of the foundation models.

#### Out of domain datasets

We used proprietary pathology datasets from Stanford, including lung and brain tumor samples. Given that some foundation models may have used CPTAC data during their pre-training, it was crucial to evaluate these models on our in-house datasets to test their generalizability and estimate their performance on real-world data. With access to detailed clinical information and molecular subtypes, we assessed these models on specific subtyping tasks, such as distinguishing between lung squamous cell carcinoma and adenocarcinoma, and differentiating between MGMT methylated and wild-type brain tumors. We also added NLST pathology^65^ and DHMC lung cancer datasets^66^ to this category as a part of our out-of-domain evaluation. This approach helped us determine the models’ true effectiveness on practical clinical tasks.

### Metadata collection for downstream tasks

For the TCGA tasks, we collected molecular subtype, gene mutation status, histologic subtype, and transcriptomic information from TCGA publications specific to the following cancer types: brain^67^, prostate^68^, lung^69,70^, breast^71^, and bladder^72^. For the pan-cancer tasks, we collected pathway information on the samples from the TCGA pan-cancer pathway study^73^ and immune groups from the TCGA immune landscape study^74^. For the CPTAC tasks, we gathered molecular subtypes and activated pathways on cases from the CPTAC pan-cancer study^56^.

### Whole Slide Image preprocessing

Whole-slide images (WSIs), initially in the SVS format, needed to be preprocessed to a machine learning-ready format. Due to the massive size of WSIs, which can often exceed dimensions of 10,000 × 10,000 pixels, it was necessary to divide them into smaller, non-overlapping tiles of 256 x 256 pixels each, at an objective magnification of 20x (0.5 microns per pixel). To accomplish this, a tissue mask was first obtained using the Otsu thresholding method^75^, which effectively separates the foreground (tissue) from the white background.

A multiple instance learning (MIL) strategy was employed, where each WSI was represented by a selection of up to 4,000 tiles. This approach was adopted to address the challenge of variable tissue density and distribution within WSIs. Tiles with excessive background or low contrast were excluded to ensure the selection of informative and representative tiles for each WSI.The selected tiles were then saved in an HDF5 database, one database per slide, to facilitate fast input/output (I/O) operations during the training and inference stages. This database organization allows for efficient access to the tiles, reducing the computational overhead associated with loading and processing large WSIs.

During the training phase, tiles were grouped into bags of *G* size. Then, in each bag, a feature vector was extracted for each tile using a given foundation model. A maximum of *G* tiles was considered in each bag, and the corresponding feature vectors were averaged. This averaged feature vector was then forwarded through a linear layer to obtain the final classification result for each WSI.This approach effectively leverages the MIL framework to model the variability within WSIs and enables the classification of slides based on the collective information extracted from their constituent tiles. The use of the HDF5 database and the efficient tile-based representation contribute to the scalability and computational efficiency of the overall pipeline.

### Vision transformers

Vision Transformers (ViTs) have significantly advanced SSL in the realm of computer vision. Originally designed for natural language processing, ViTs adapt the transformer architecture to visual tasks by dividing images into smaller patches treated as tokens. The self-attention mechanisms in ViTs enable the model to capture intricate details and long-range dependencies within images, providing a superior contextual understanding compared to traditional convolutional neural network-based methods, especially in complex domains like pathology ^76^.

### Foundation models

In the past couple of years, several ViT based pathology models like Lunit-Dino^24^, Phikon(Owkin)^33^, UNI^25^, Virchow^26^ have been trained on huge datasets on multiple tissue types and evaluated on a variety of downstream tasks. We evaluated in-domain foundation models (i.e., models trained on digital pathology data such as Lunit-Dino), as well as out-of-domain vision models, trained on natural images (ImageNet data) such as DINO from Meta AI. We further categorized these models as general vision only (natural image based), general vision-language (natural images with text), pathology vision models (digital pathology image-based) and pathology vision-language models (pathology images and medical text). We conducted a comparative analysis across the different model categories to assess their relative performance. The pathology domain-specific models have been primarily trained on publicly available datasets like TCGA, and/or on proprietary datasets (for example, in the case of UNI). These foundation models are built upon ViT-small, ViT-base, and ViT-large/ViT-huge architectures as their core backbones. In our work, we benchmark these models using identical tasks, datasets, and hyperparameters.

### Computational resources

For our evaluation experiments, we used the Polaris supercomputer at the Argonne National Laboratory and the Stanford University Sherlock cluster (Stanford Research Computing Center).

### Model Evaluation

We assessed the performance of these models by applying a linear classifier to frozen extracted features, using consistent evaluation settings across all models. We froze the pretrained model and extracted features for the downstream task of interest, before doing linear probing. For our evaluation protocol, we used the same hyperparameters across all models and tasks and did not perform any data augmentation. For linear evaluation, we used a learning rate of 0.001, batch size of 16 per gpu, and trained the linear layer for 30 epochs. All tasks were run on a single node equipped with four NVIDIA A100 GPUs, each with 40 GB of memory. The foundation models varied based on different architecture sizes like ViT-S, ViT-B, ViT-L as well as patch sizes. We divided datasets for TCGA, CPTAC and out-of-domain evaluation downstream tasks into training, validation and test sets with a 70/15/15 split. We also implemented 5-fold cross-validation, complemented by evaluation on a separate independent test set, to robustly assess model performance for these tasks. We report the performance of models on independent test sets in our study. For external public benchmarking tasks, the datasets are usually available as training and test set or validation set. We report the performance on validation or test sets as per their availability. We also evaluated these models on slide level tasks from TCGA, CPTAC and out of domain evaluation datasets as well as patch based classification tasks from external benchmarking tasks separately. Performance is evaluated using metrics including balanced accuracy, precision, recall, and F1 score. We computed the arithmetic mean of the four metrics to derive the ‘average performance’ providing a balanced summary of overall model effectiveness.

## Data Availability

All data produced in the present work is available at a dashboard

https://pathbench.stanford.edu/

## Acknowledgements

Research reported here was further supported by the National Cancer Institute (NCI) under awards: R01 CA260271. The content is solely the responsibility of the authors and does not necessarily represent the official views of the National Institutes of Health. This research was further supported by the Argonne Leadership Computing Facility, a U.S. Department of Energy (DOE) Office of Science user facility at Argonne National Laboratory.

## Supplementary Figure Legends

**Supplementary Figure 1.**
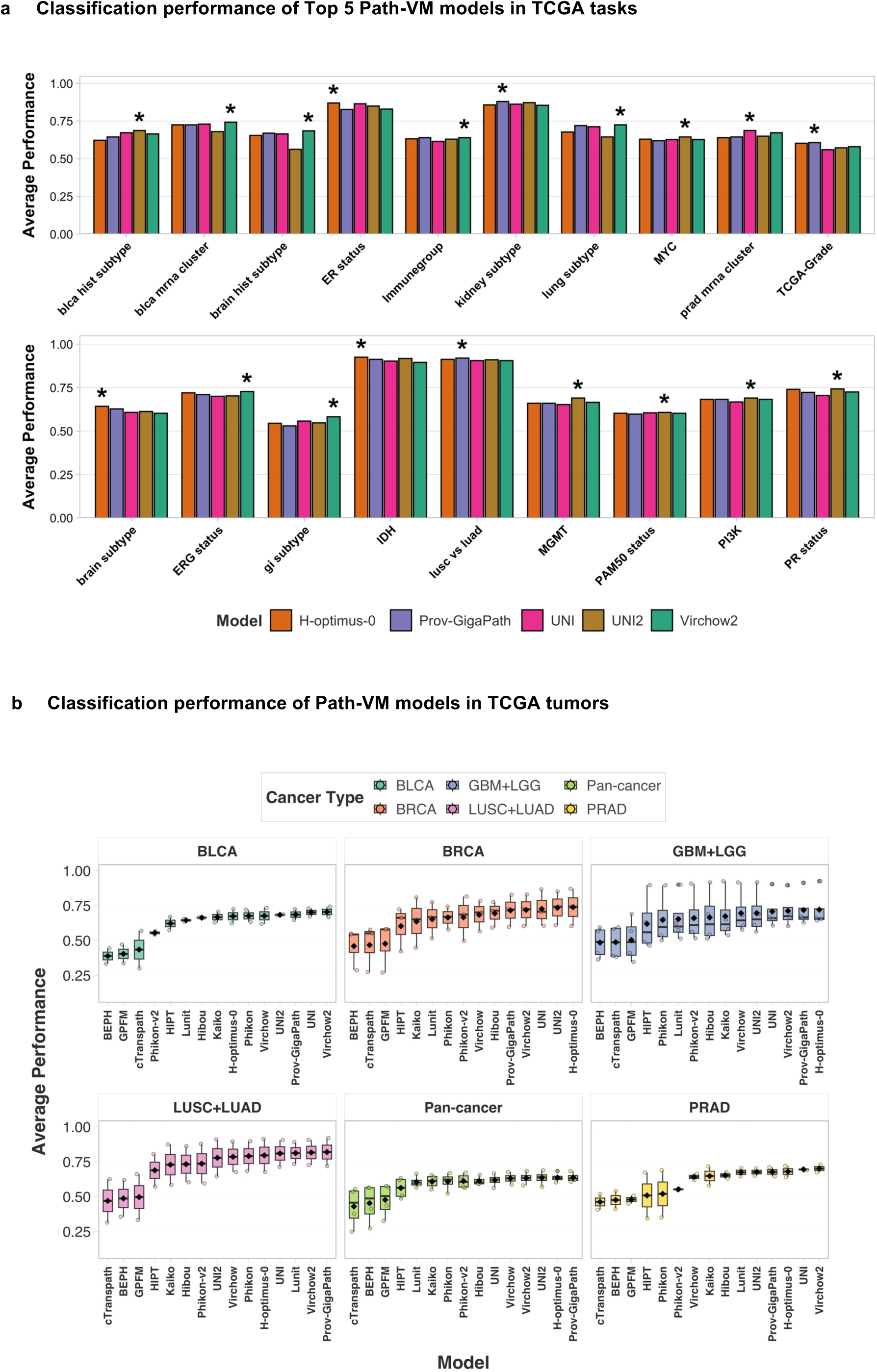
Classification performance of pathology-specific vision models (Path-VM) on TCGA tasks. **(a)** Top 5 performing Path-VM in each TCGA task. **(b)** Model-wise classification performance across individual TCGA tumor types, sorted by average performance per model. This figure highlights the relative strengths of each model across different tumor types and underscores variability in model generalization.

**Supplementary Figure 2.**
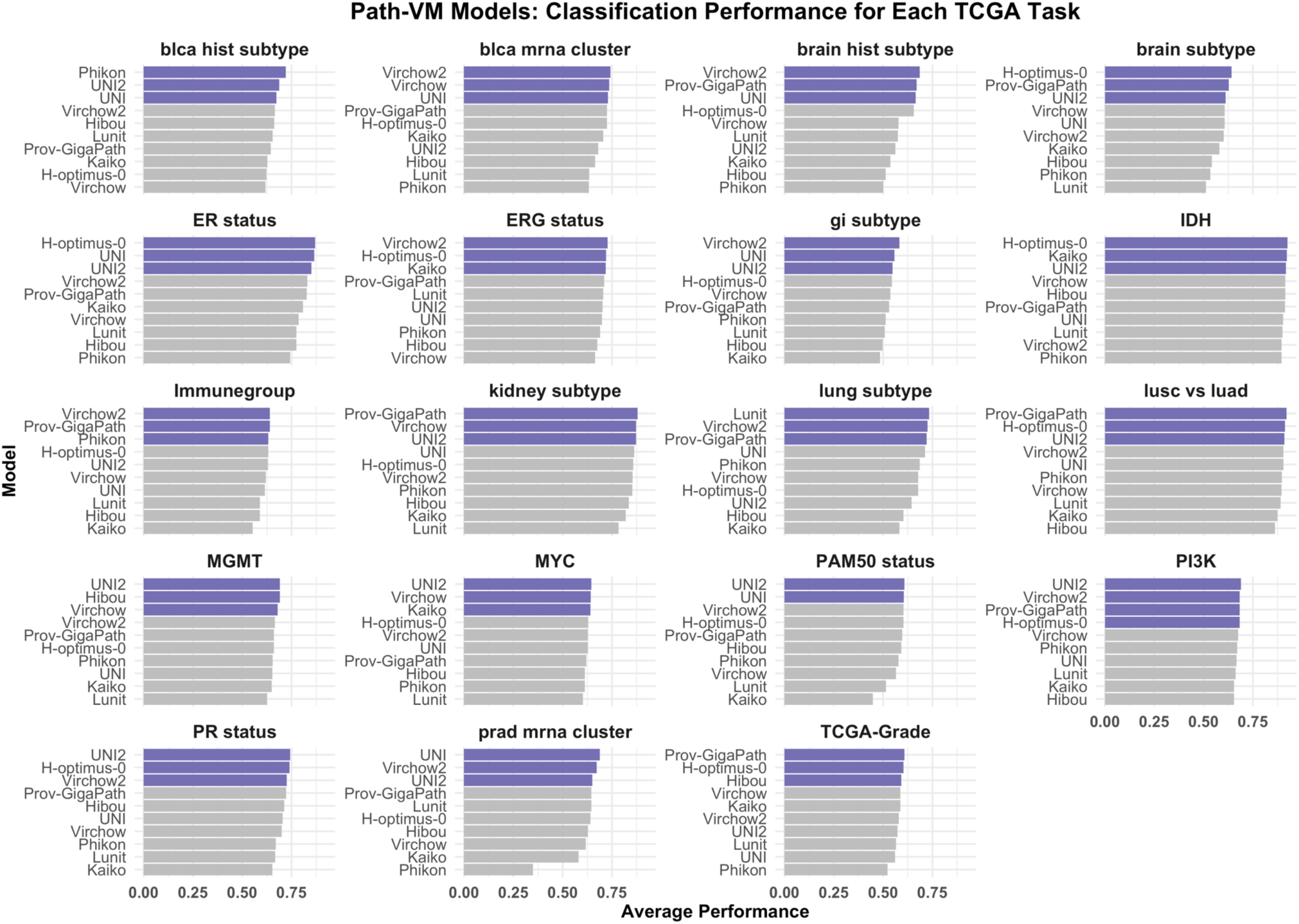
Classification performance of top 10 pathology-specific vision models (Path-VM) on each TCGA task. (a) Top 10 performing Path-VM in each TCGA task, ranked by average performance. The top three models are highlighted in purple for emphasis.

**Supplementary Figure 3.**
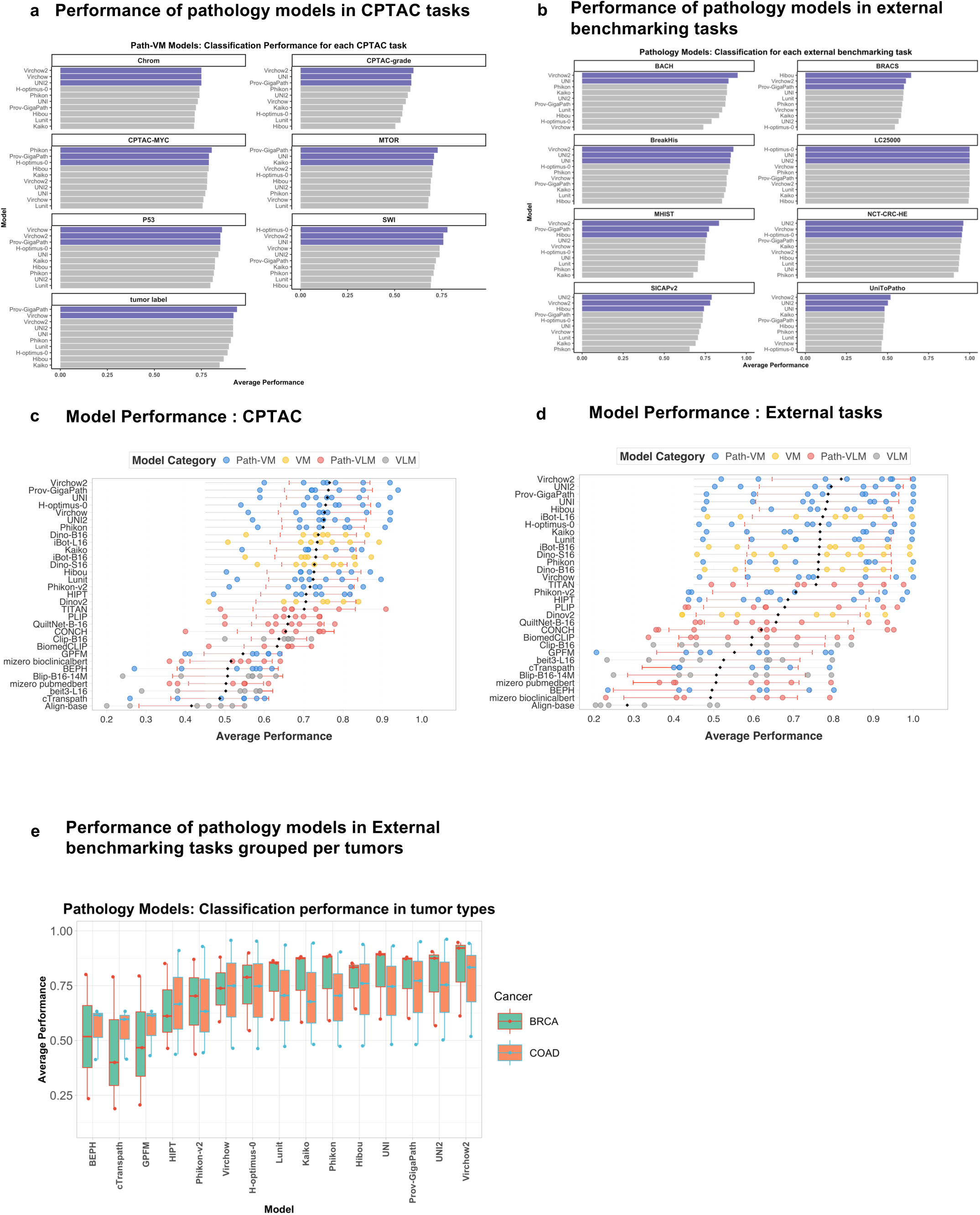
Classification performance in CPTAC and external benchmarking tasks. (a) Top 10 performing pathology-specific vision models (Path-VM) in each CPTAC task, ranked by average performance. The top three models are highlighted in purple for emphasis. (b) Top 10 performing Path-VM in each external task, ranked by average performance, with the top three models highlighted in purple for emphasis. (c) Performance of all 31 models across, sorted by mean of average performance across CPTAC tasks. (d) Performance of all 31 models, sorted by mean of average performance, across external tasks. (e) Performance of Path-VM models on external tasks, grouped by tumor types BRCA and COAD.

**Supplementary Figure 4.**
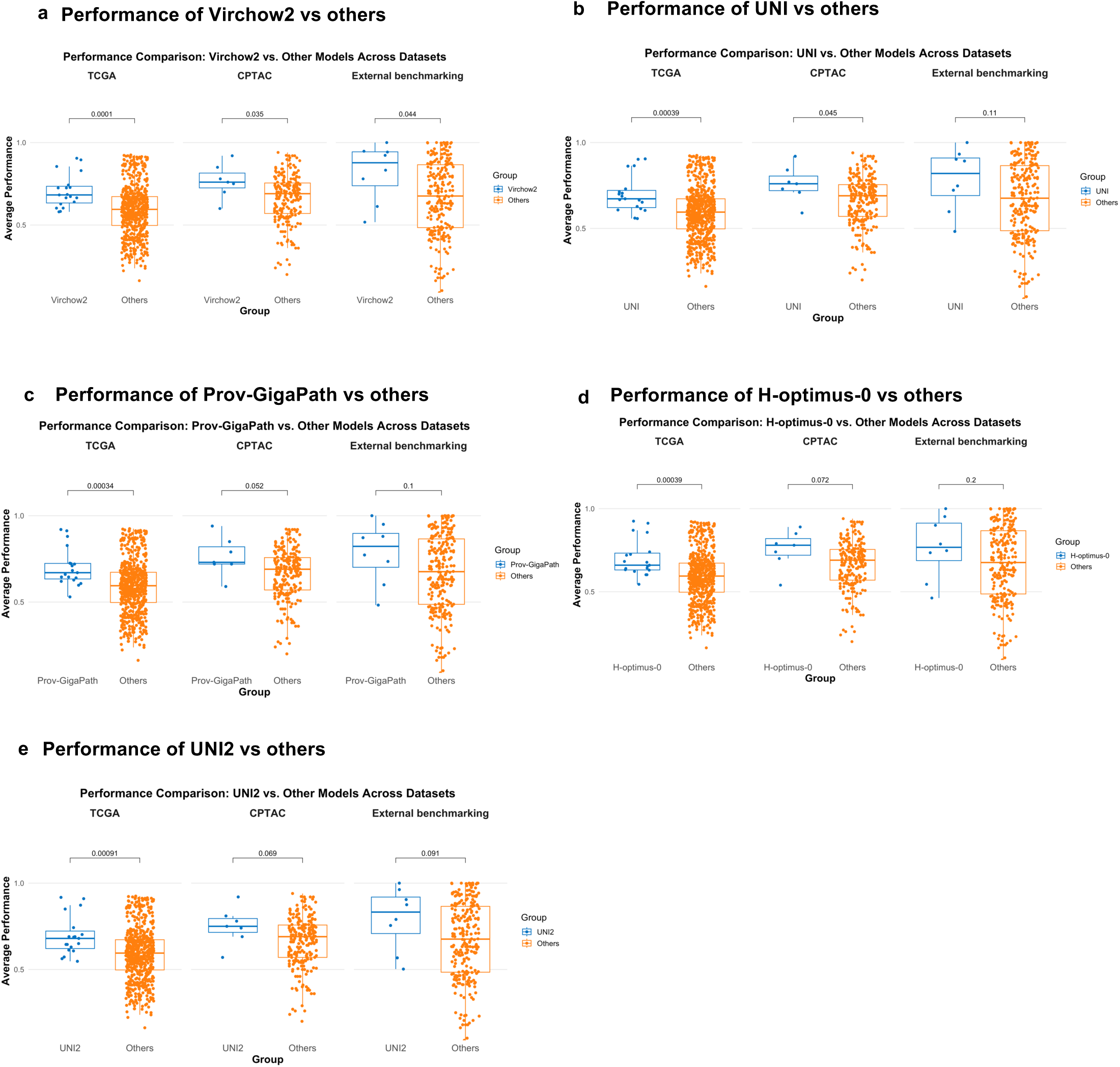
Classification Performance of top pathology-specific vision(Path-VM) models vs other models. (a) Performance comparison between Virchow2 vs rest of other models. (b) Performance comparison between UNI vs rest of other models. (c) Performance comparison between Prov-GigaPath vs rest of other models. (d) Performance comparison between H-optimus-0 vs rest of other models. (e) Performance comparison between UNI2 vs rest of other models. Significance levels: * p < 0.05, ** p < 0.01, *** p < 0.001 (t-tests with Bonferroni correction)

**Supplementary Figure 5.**
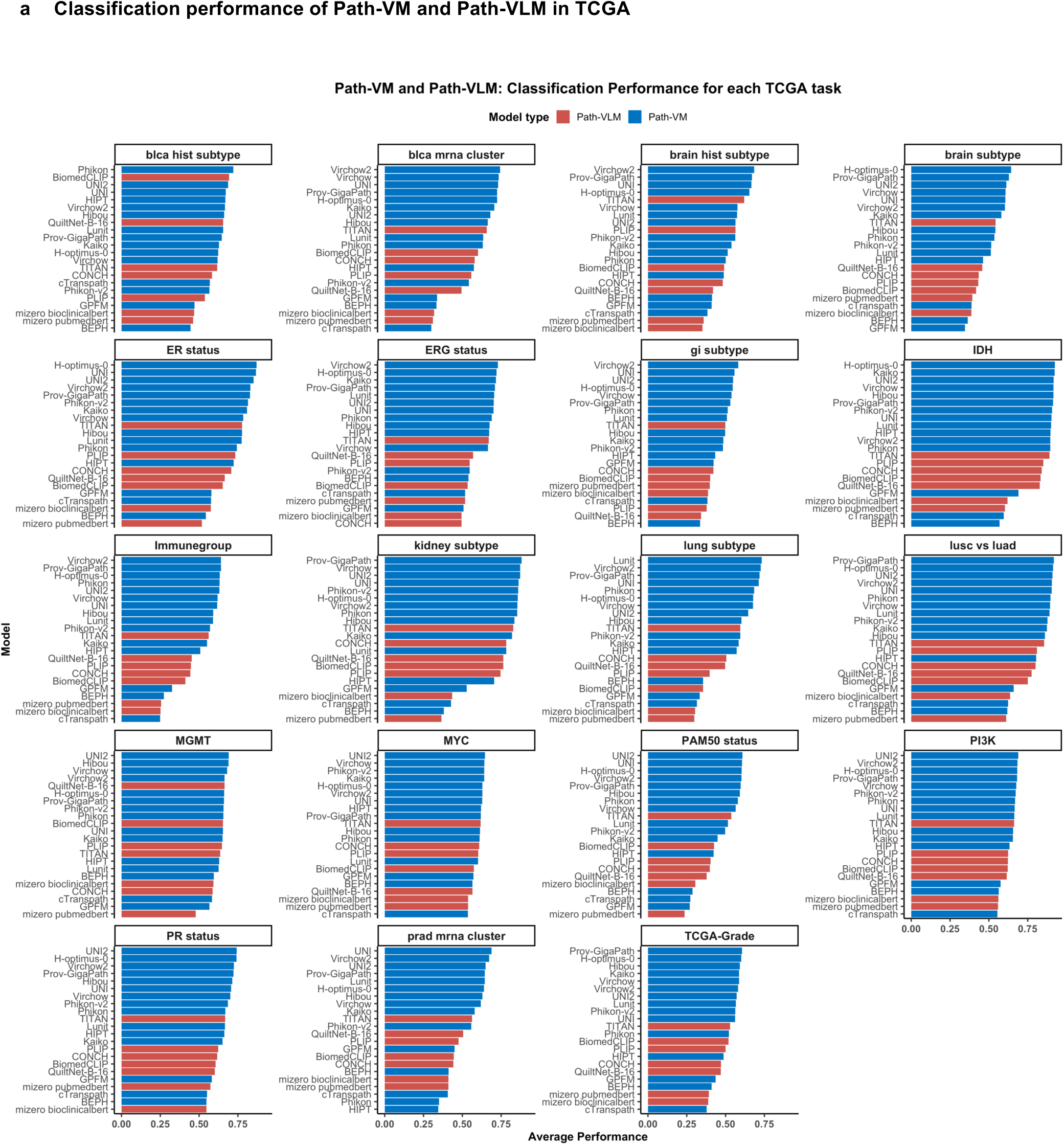
Classification performance of pathology-specific vision models (Path-VM)and pathology-specific vision-language models (Path-VLM) in TCGA tasks. (a) Performance of all Path-VM and Path-VLM in each TCGA task, ranked by average performance, with the top three models highlighted in pink for emphasis.

**Supplementary Figure 6.**
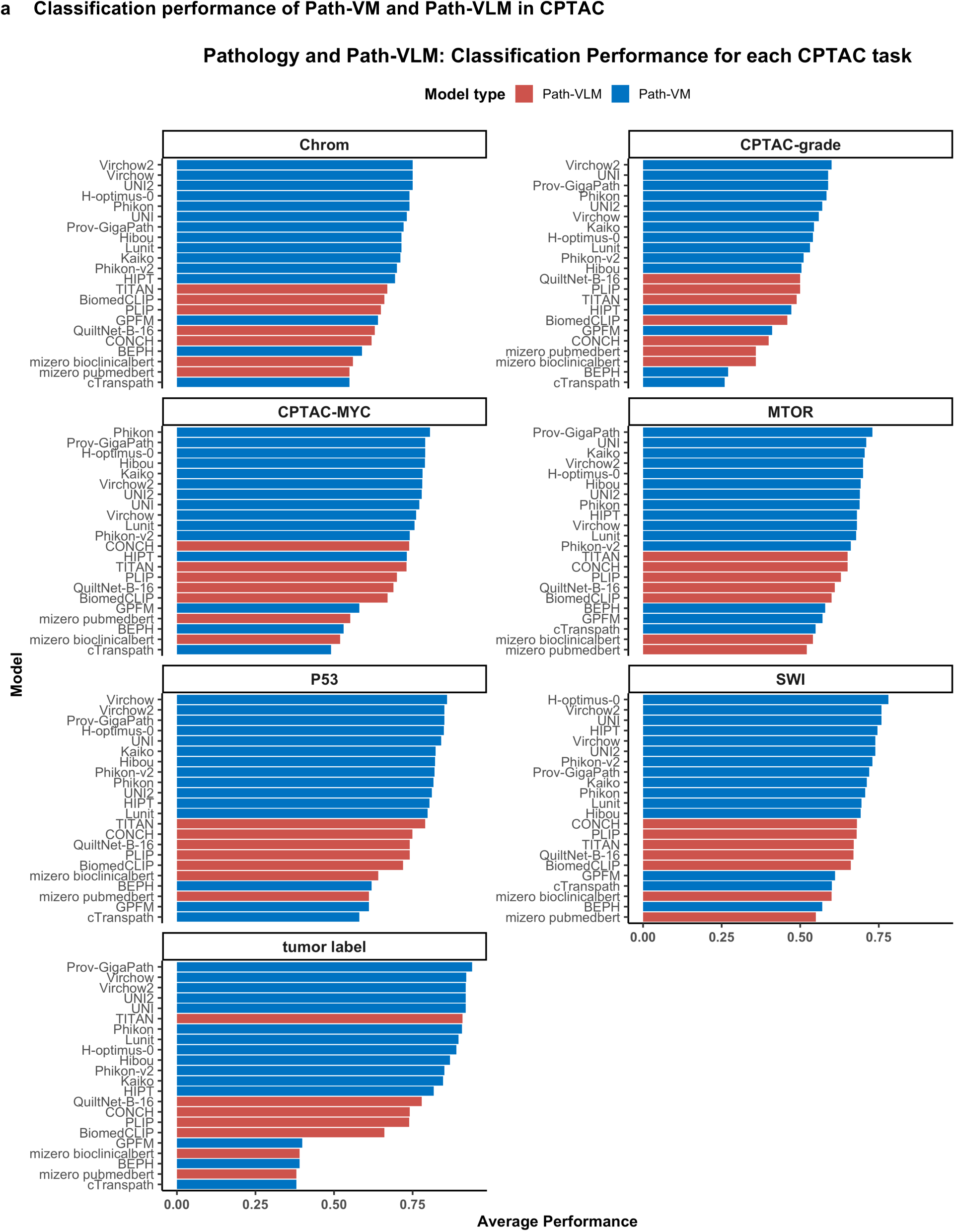
Classification performance of pathology-specific vision models (Path-VM)and pathology-specific vision-language models (Path-VLM) in CPTAC tasks. (a) Performance of all Path-VM and Path-VLM in each CPTAC task, ranked by average performance, with the top three models highlighted in pink for emphasis.

**Supplementary Figure 7.**
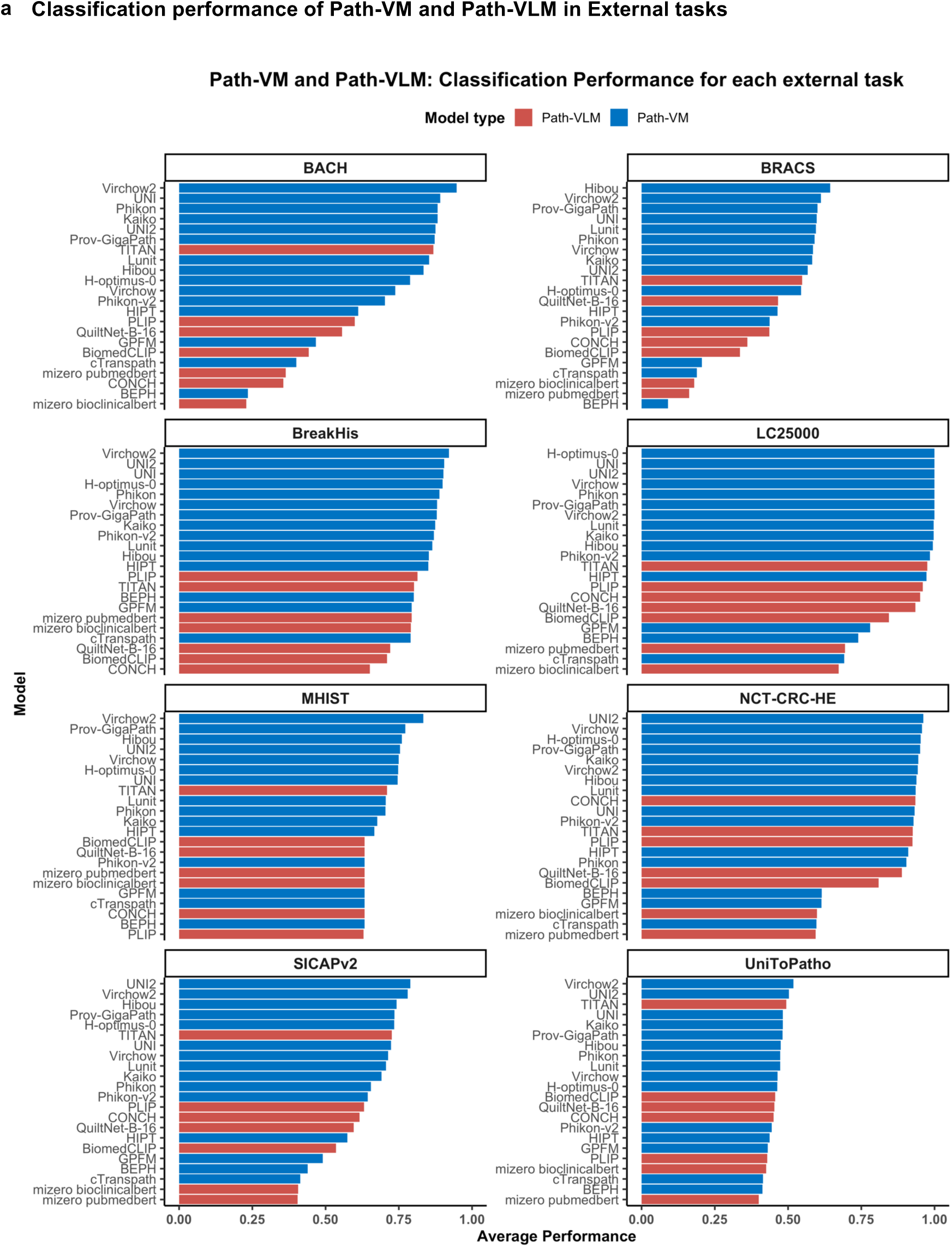
Classification performance of pathology-specific vision models (Path-VM)and pathology-specific vision-language models (Path-VLM) in external tasks. (a) Performance of all Path-VM and Path-VLM in each external task, ranked by average performance, with the top three models highlighted in pink for emphasis.

**Supplementary Figure 8.**
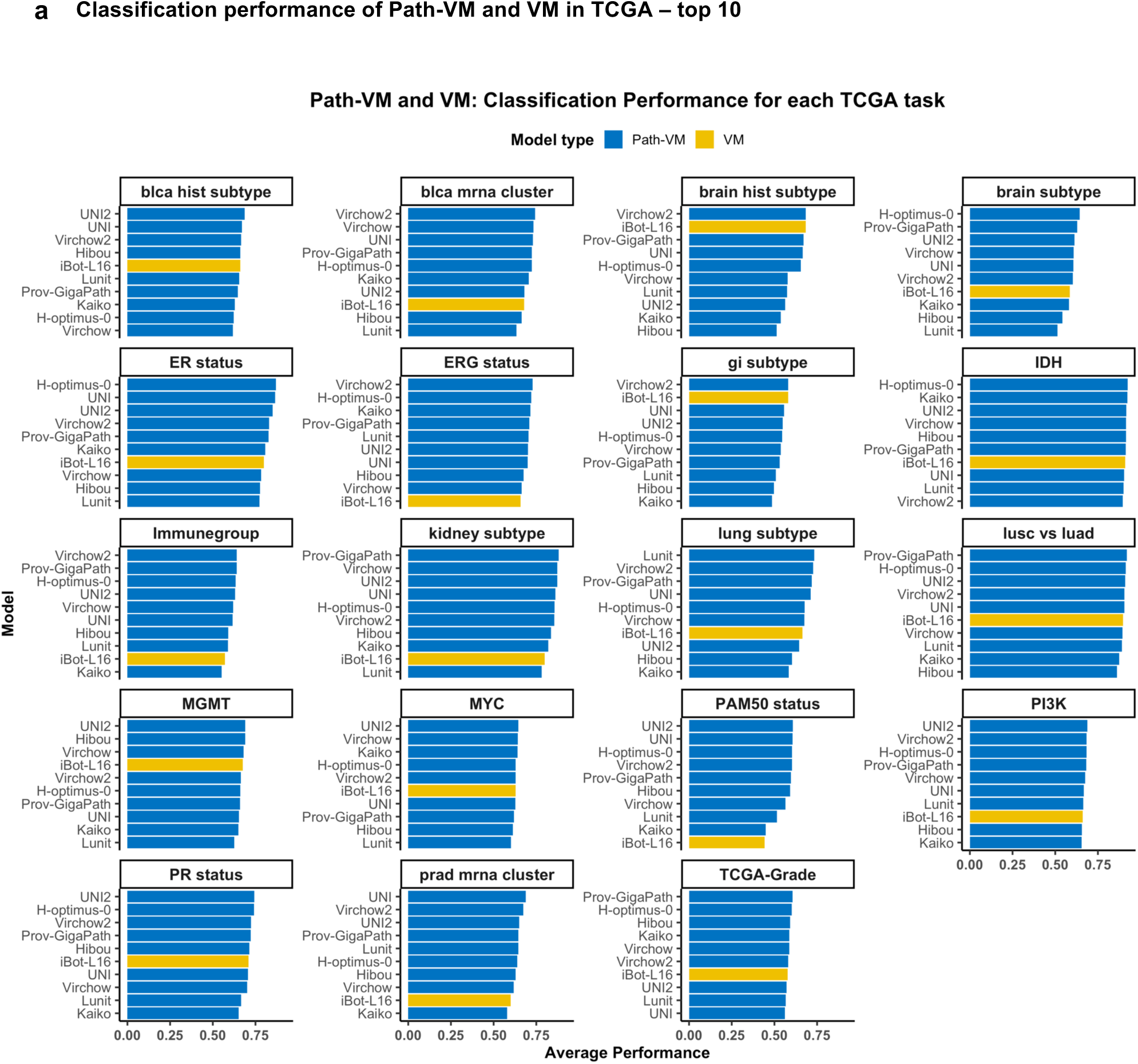
Classification performance of pathology-specific vision models (Path-VM) and general vision models (VM) in TCGA tasks. (a) Top 10 performing pathology and vision models in each TCGA task, ranked by average performance, with the top three models visually emphasized in pink.

**Supplementary Figure 9.**
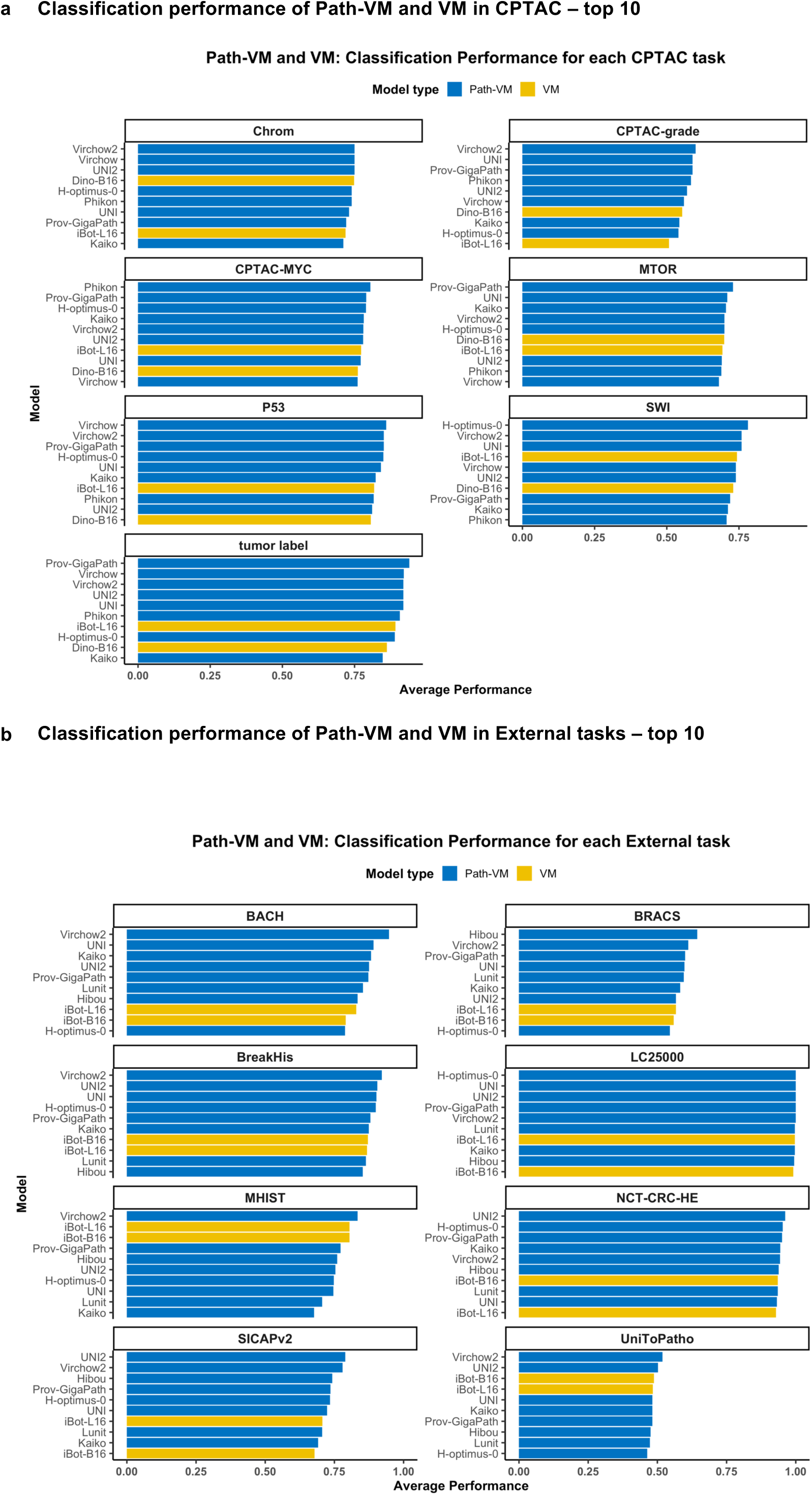
Classification performance of pathology-specific vision and general vision models in CPTAC and external tasks. (a) Top 10 performing Path-VM and VM in each CPTAC task, ranked by average performance, with the top three models visually emphasized in pink. (b) Top 10 performing Path-VM and VM in each external task, ranked by average performance, with the top three models visually emphasized in pink.

**Supplementary Figure 10.**
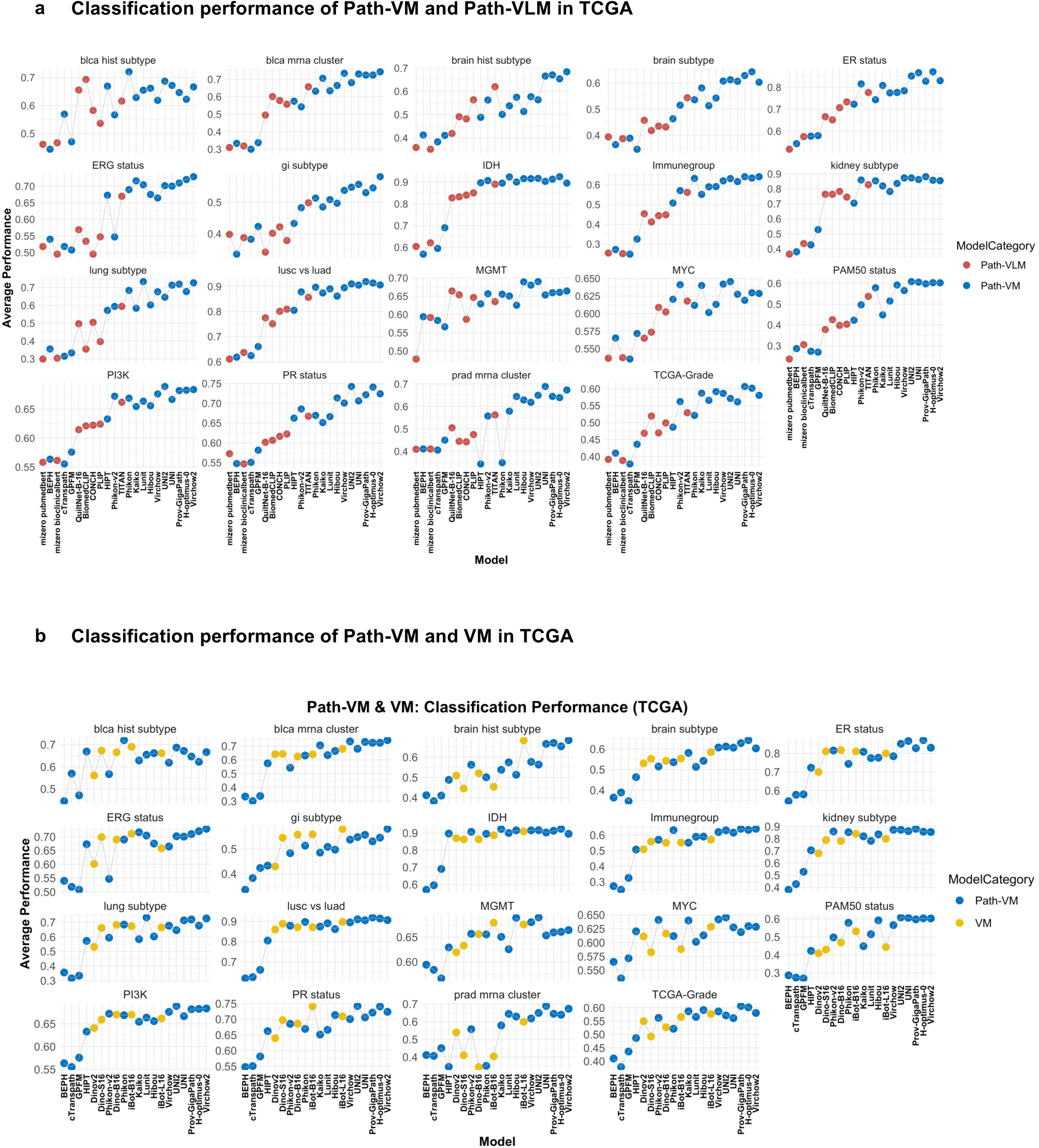
Classification performance in TCGA. (a) Performance of all Path-VM and Path-VLM in each TCGA task, based on average performance (b) Performance of all Path-VM and VM in each TCGA task, based on average performance.

**Supplementary Figure 11.**
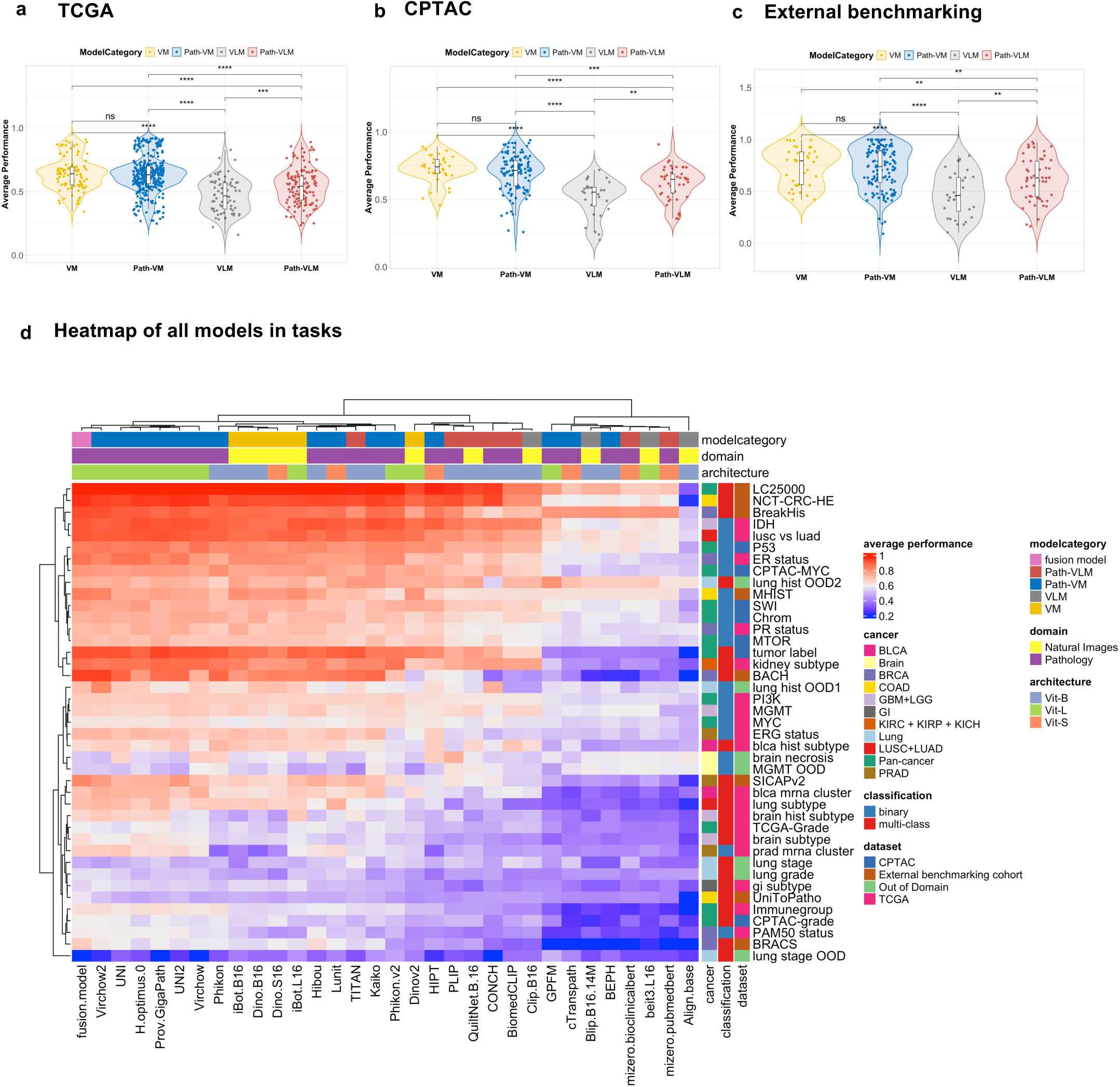
Comparison between model categories. Performance comparison of general vision, pathology-specific vision models, general vision language models and pathology-specific vision-language models in (a) TCGA (b) CPTAC (c) External benchmarking tasks. Significance levels: * p < 0.05, ** p < 0.01, *** p < 0.001 (pairwise t-tests with Bonferroni correction) (d) Heatmap of performance of all models in all tasks including out of domain tasks.

**Supplementary Figure 12.**
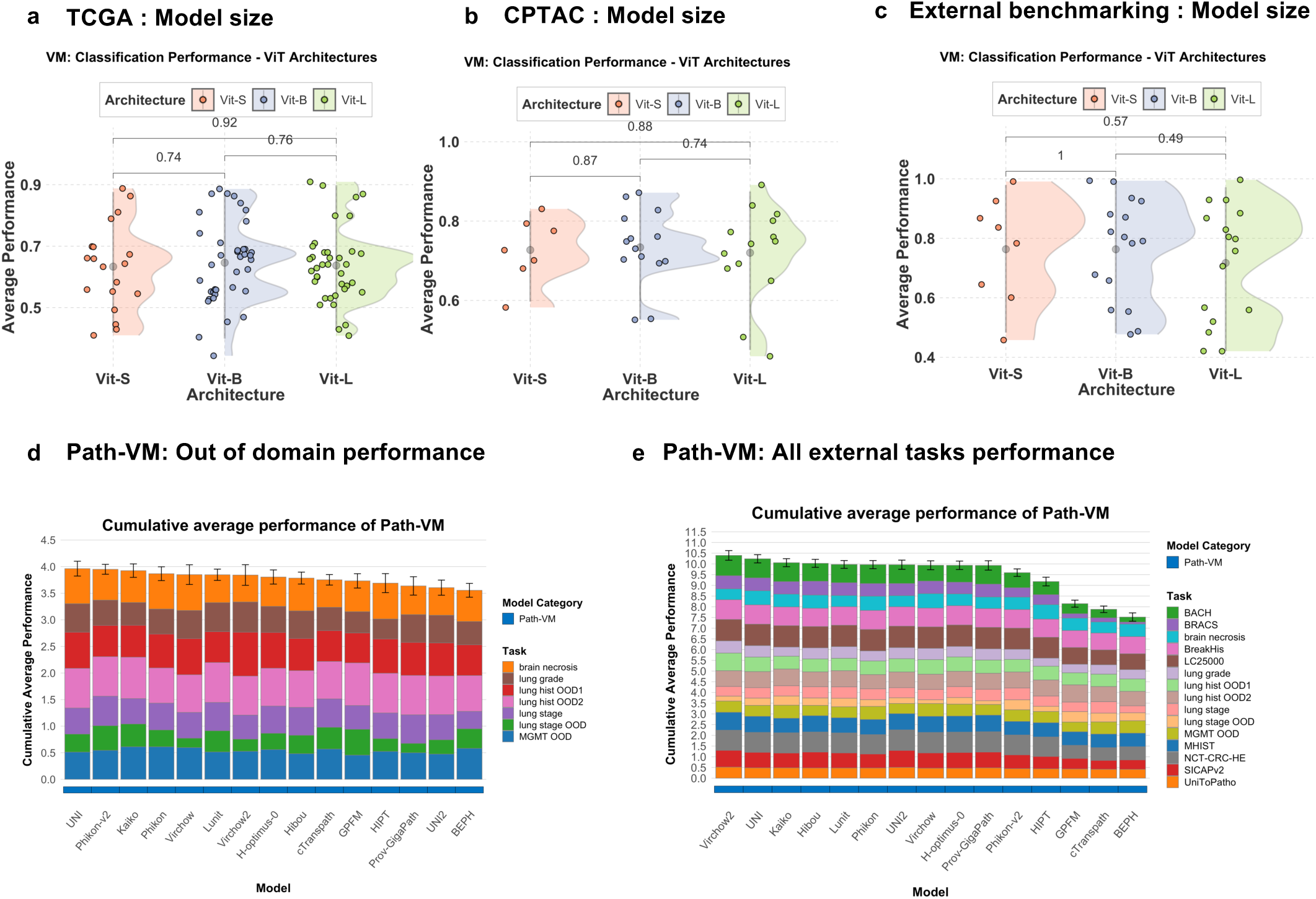
Comparison between Model Architectures and performance of pathology models in external tasks. Performance comparison of vision model architectures ViT-S, Vit-B, ViT-L in (a) TCGA (b) CPTAC (c) External benchmarking tasks (d) Cumulative average performance of Path-VM across out of domain tasks, sorted in descending order. (e) Cumulative average performance of Path-VM across external benchmarking tasks and out of domain tasks, sorted in descending order.

